# A Time-dependent mathematical model for COVID-19 transmission dynamics and analysis of critical and hospitalized cases with bed requirements

**DOI:** 10.1101/2020.10.28.20221721

**Authors:** Avaneesh Singh, Manish Kumar Bajpai, Shyam Lal Gupta

**Author notes:** Corresponding Author: Avaneesh Singh.

## Abstract

A time-dependent SEAIHCRD model is the extension of the SEIR model, which includes some new compartment that is asymptomatic infectious people, hospitalized people, critical people, and dead compartments. In this article, we analyzed six countries, namely the United States, Brazil, India, South Africa, Russia, and Mexico. A time-dependent SEAIHCRD model calculates the magnitude of peaks for exposed people, asymptomatic infectious people, symptomatic infectious people, hospitalized people, the number of people admitted to ICUs, and the number of COVID-19 deaths over time. It also computes the spread scenario and endpoints of disease. The proposed model also involves asymptomatic infectious individuals. To estimate the various parameters, we first collect the data and fit that using the Lavenberg-Marquardt model for death cases. Then we calculate infection rate, recovery rate, case fatality rate, and the basic reproduction number over time. We calculate two types of case fatality rates: one is the daily case fatality rate, and the other is the total case fatality rate. The proposed model includes the social distance parameter, various age classes, hospital beds for severe cases, and ICU beds or ventilators for critical cases. This model will be useful to determine various essential parameters such as daily hospitalization rate, daily death rates, including the requirement of normal and ICU beds during peak days of infection.

## 1 Introduction

Many countries have suffered a lot from this COVID**-**19. Some countries suffer from loss of beds and ICU beds. Due to triage conditions, doctors have to select critical conditioned people for treatment, and some patients have not been treated due to a lack of hospital beds and ICU beds. It is regrettable to us that someone dies without treatment. Perhaps if they were treated, then save their lives. If the government has prior knowledge of how much beds are needed in a particular time, then the arrangement can be made, and it can save many lives for those who have died of lack of facility. Our proposed model is a new mathematical method that extends the SEIR model by adding death, hospitalized, and critical compartments. The hospitalized compartment and critical compartment are to a new compartment added in this model to enhance the disease transmission visibility. No one has used these two compartments in any model of my knowledge until that time. The model also calculates how many hospital beds and ICU beds are needed in peak time. The proposed model also calculates the case fatality rate and the basic reproduction rate.The calculation process for requiring beds and ICU beds by this method is new. We calculate the basic reproduction number using the logistic regression over time; its unique idea is used in this model. We also consider age groups for case fatality rate analysis; if any country’s elderly population is more, then the case fatality rate will be higher in that country.

The entire world is combating against a new enemy in these days, which is the COVID-19 virus. The virus has spread exponentially around the world since its first appearance in China. Most of the countries are struggling with economic and social fronts due to this virus. Tyrell and Bynoe described the coronavirus for the first time in 1966 [1]. An outbreak of pneumonia was reported in Wuhan, China, in December 2019 [2]. The virus causing that outbreak was provisionally named as 2019-nCoV, which was renames as SARS-CoV-2 by the International Committee on Virus Taxonomy [3-5]. The World Health Organization announced these outbreaks as an international public health emergency on January 30 and a pandemic on March 11, 2020 [6, 7]. COVID-19 caused a global recession and has an impact on social life [8]. More than 22.4 million cases, out of which more than 14.3 million people have recovered and around 788,000 have died, of COVID-19 pandemic have been reported across 188 countries and territories till August 20, 2020. [9]. Worldwide, researchers are working 24 hours a day to find the SARS-CoV-2 virus vaccine. There are nearly 231 vaccines reported as potential candidates for the cure of COVID-19 till August 2020; however, none of these have successfully established their safety and efficacy in clinical trials. As per various reports, nearly 25 vaccine candidates are in different stages of clinical trials [10-12].

Asymptomatic cases mean people without symptoms. However, there are reports of loss of sense of smell in people who otherwise have no symptoms. This is now recognized as an official symptom of the virus, and people experiencing it should self-isolate themselves. Reports from many countries suggest that asymptomatic infected individuals are much less likely to transmit the virus than those with symptoms [13]. Most of the symptomatic cases are mild. They do not have to be in the hospital. They can be quarantined at home under the guidance of the doctor. Temperature with minor cough, tiredness, moderate breathlessness, pain in the muscles, headache, sour throat, and diarrhea are the main signs of mild cases. Symptoms of severe cases are more visible in older people. Its symptoms are rarely seen in healthy young people. The main symptoms of severe cases are pneumonia that involves excessive breathlessness, chest pain, high temperature, low oxygen levels, rapid heart rate, and decreased blood pressure. Severe cases need hospital beds for the treatment of patients. Symptoms of critical cases are more visible in older people who have pre-existing diseases such as cardiovascular disease, diabetes, chronic respiratory disease, hypertension, and cancer. Many organs fail to function because of this. Critical cases need ventilators or ICU care for the treatment of patients. COVID-19 shows an increased number of cases and a greater risk of severe disease with increasing age. [14, 15]

Mathematical models that forecast the possible effects of various approaches to behavioural changes in the population are an essential source of knowledge for policymakers [16, 17]. Currently, the countries most affected by COVID-19 disease in the world are the United States, Brazil, India, South Africa, Russia, and Mexico. In the present work, we are introducing a new mathematical model for the study of the Pandemic Transmission and Control Dynamics of COVID-19 based on the reports of these affected countries. Compartmental models are composed of ordinary differential equations. These models are separated into many compartments. Our proposed SEAIHCRD model has eight compartments: susceptible (S), exposed (E), asymptomatic infectious (A), symptomatic infectious (I), hospitalized (H), critical (C), recovered (R), and deceased or death (D), collectively termed as SEAIHCRD. The basic rule of the compartmental model is the transition of people from one compartment to another. Models use mathematics to find the different parameters to estimate the effect of various interventions and then to decide which intervention to stop and which to continue. These models are primarily used in epidemiology and many other fields, such as economics, politics, social science, and medicine, etc.

Kermack and Mckendrick have introduced the first basic infectious disease compartmental model, SIR [18]. The SEIR model is a commonly used epidemiological model based on the SIR model [19]. The Kermack and Mckendrick epidemic model has predicted outbreak behaviour very close to the real scenarios seen in many recorded epidemics [19]. Since Kermack and Mckendrick’s paper, stochastic models, discrete-time models, continuous-time models, and diffusion models have been developed for many diseases. Some manuscripts provide an excellent approach to mathematical models that provide a comprehensive understanding of the disease outbreak scenario [20-34]. The SEIR model has not been able to estimate spread where preventive measures such as social distances, different age groups, number of ICU beds, number of hospital beds, and mortality rates have been adopted. The proposed model in this work is a modification to the SEIR model. The present manuscript includes an updated SEIR model for COVID-19 with all the above parameters. We propose a prediction model that has been constrained by many observed data under uncertainties. The time-dependent SEAIHCRD model estimates the magnitude of peaks for exposed people, the number of asymptomatic infectious people, the number of symptomatic infectious people, the number of people hospitalized, the number of people admitted to ICUs and the number of deaths of COVID-19 over time. The proposed method includes asymptomatic infectious cases as a separate compartment. Asymptomatic infection cases are either recovered or, if symptoms begin to appear within a few days, enter into the symptomatic infectious compartment.

The time-dependent SEAIHCRD model includes a social distancing parameter, analysis of different age groups, number of ICU beds, number of hospital beds, and lock-down effect, which gives a more accurate estimate of the future requirement of the beds normal as well as the ICU in the hospitals. The proposed model also analyzes the basic reproduction number over time. Early stages of the outbreak, the basic reproductive number (R0) in January ranged from 1.4 to 2.5, but a subsequent study suggested that it may be around 5.7 [35, 36]. The spread of disease in these areas has been stable or declining since mid-May 2020[37]. The number of cases doubled in almost seven and a half days in the early stages of the outbreak, but later the rate increased [38]. The proposed model analyzes the case fatality rate over time. The case fatality rate is the ratio of people dying from a specific disease among all individuals suffering from the disease over a certain period. [39, 40].

The structure of this paper is as follows; Section 1 introduces COVID-19 and explains the significance of this research. A time-dependent SEAIHCRD predictive mathematical model for COVID-19 is presented in section 2. Section 3 discusses the data set and its analysis used to validate the proposed mathematical models. Results and discussions on the proposed model are discussed in Section 4. Finally, the conclusion and some future works are discussed in Section 5.

## 2. Proposed Methodology

Modelling is one of the most powerful tools that give intuitive inferences when multiple factors act together. Compartment models such as SIR and SEIR are mostly used today for the mathematical modelling of infectious diseases. Over the last few weeks, many epidemiologists and data scientists have worked on mathematical modelling of infectious diseases, in particular, Coronavirus disease (COVID-19). Coronavirus disease (COVID-19) is an infectious disease that can spread from one member of the population to another. Kermack-McKendrick [18] proposed the basic compartmental SIR model in 1927. Beretta and Takeuchi [41] have developed their theories based on research conducted by Kermack and McKendrick [18] and have proposed a SIR model on time delays. In the spread of infectious diseases, Cooke and Driessche [42] studied the incubation cycle, developed the “Exposed, E” compartment, and suggested a time delay SEIR model.

Singh A et al. (2020) described the basic compartment model SIR, the SEIR model, and the SEIRD model [27]. Our proposed model, a time-dependent SEAIHCRD model, is a new mathematical method that extends the SEIR model by adding asymptomatic infectious, death, hospitalized, and critical compartments.

COVID-19 Infectious disease is essentially categorized as symptomatic infectious and asymptomatic infectious cases. Fig.1 shows the categorization of infectious cases and the flow of infectious cases. There is no need to hospitalize asymptomatic cases and mild cases to reduce the pressure on hospitals. At the same time, it would ensure that serious and urgent patients have access to beds when they are most needed. There may be fear among the positive patients that they are ignored and not provided the necessary treatment. However, there is no need for fear, as mild cases can be cured even if the patients stay at home in isolation with the guidelines as prescribed by health professionals [44].

**Figure 1:**
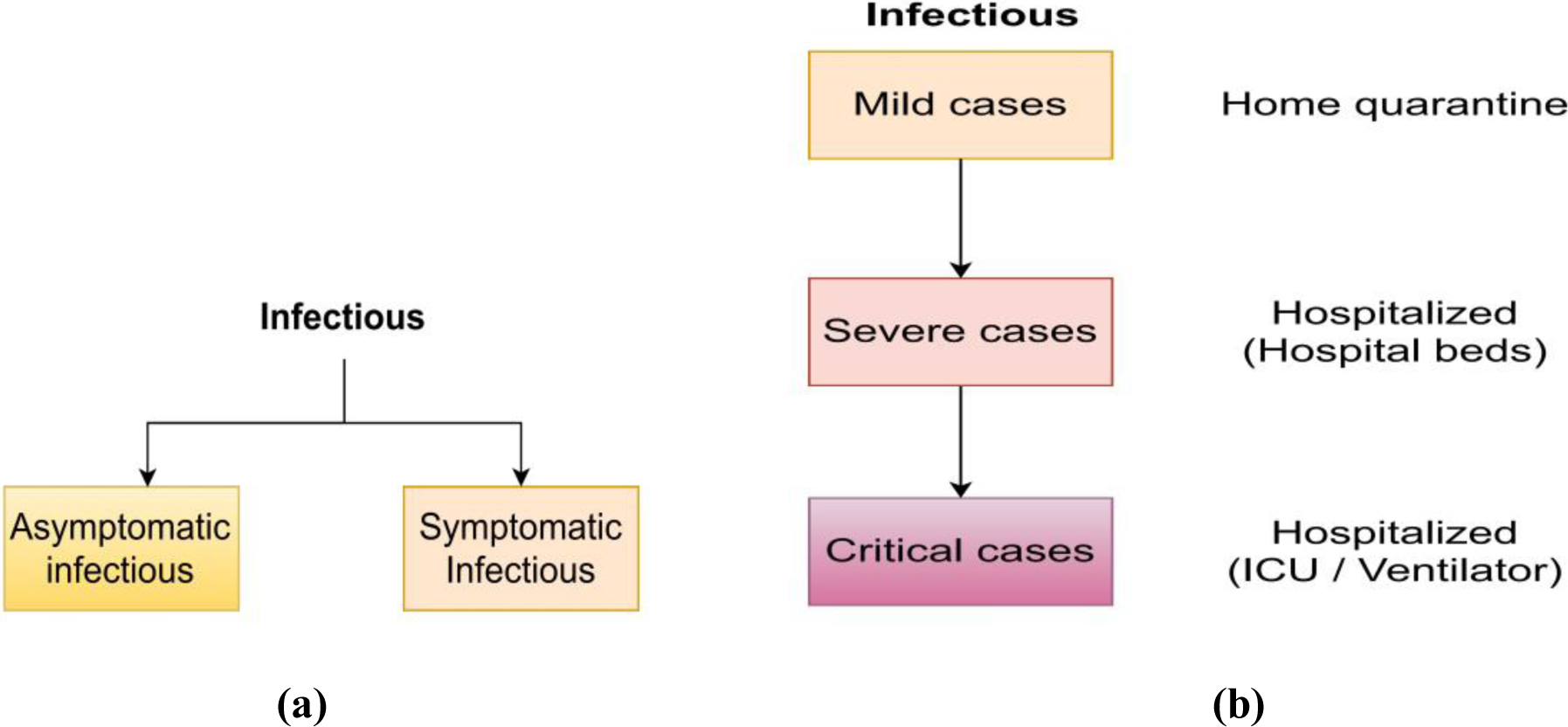
**(a)** Infectious categorization, **(b)** Infectious cases flow

In Fig. 2, the Actor is a source of infection, and it transfers the infection from one Actor to another. The Actor may be either symptomatic or asymptomatic infected. Most of the cases are mild or asymptomatic. Severe cases are significantly less than mild cases but more than critical cases. Initially, some cases are mild, but with time, it becomes severe or critical; then, they need hospital beds or ICU beds.

**Figure 2:**
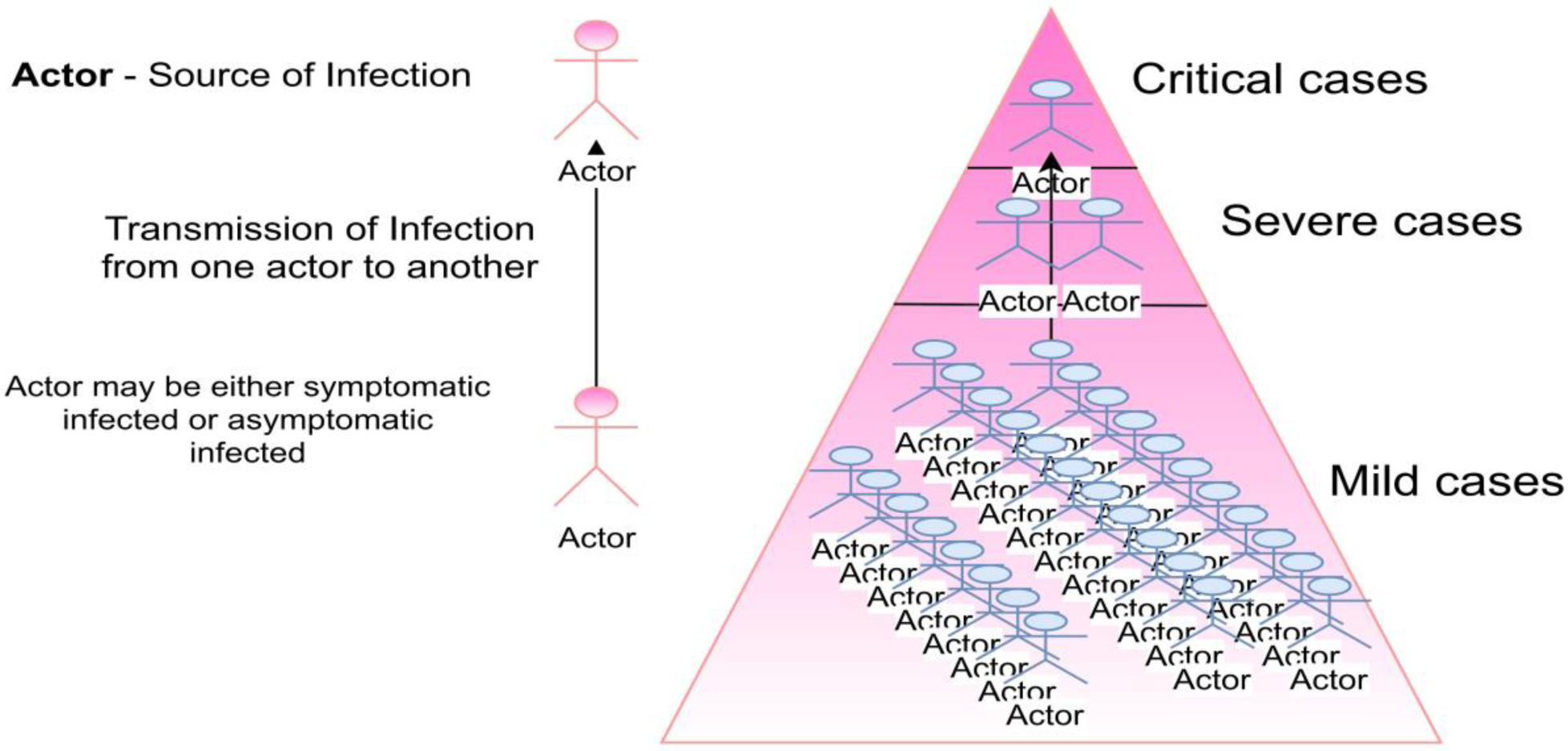
Hierarchy of COVID-19 Cases in numbers

Most cases are asymptomatic because many cases are missing from the symptom-based screening test. Children and young adults may become asymptomatic. ICMR has advised asymptomatic and COVID positive cases with mild symptoms like fever and cough to observe self-isolation and put under home quarantine for at least 17 days; hence the pressure on the designate COVID hospitals is reduced [43-44].

The non-infected and asymptomatic population is susceptible to COVID-19. There is an incubation period for the susceptible, exposed person (compartment E), which is defined as the duration in which one exposed may become infected with symptoms or asymptomatic. Asymptomatic infected individuals can enter the Symptomatic infected or recover. From Symptomatic infected, they can either die or recover shown in Fig. 3.

**Figure 3:**
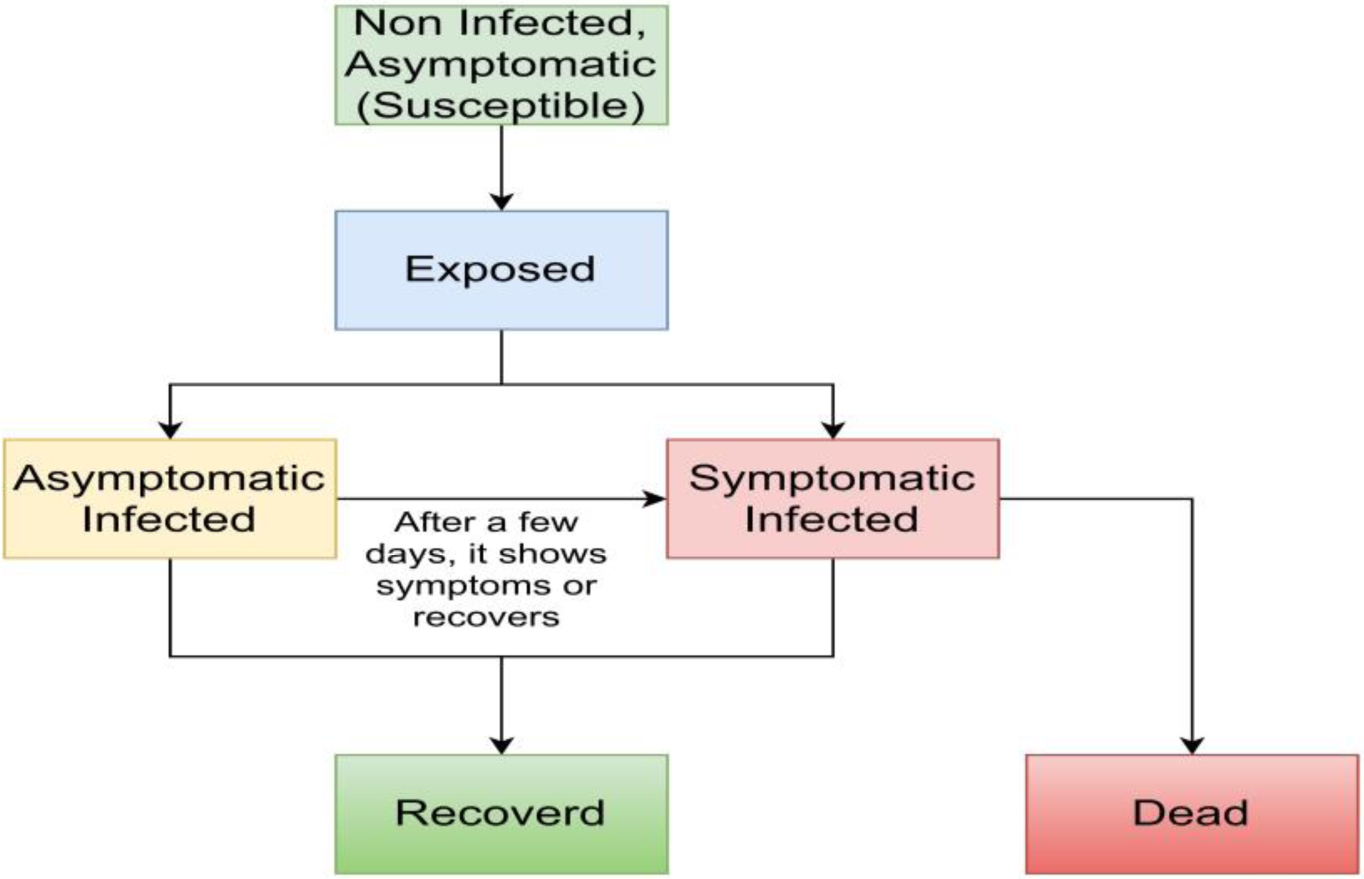
Flow of Asymptomatic infectious cases

### 2.1 A time-dependent SEAIHCRD model

In the present manuscript, the Asymptomatic Infectious, Hospitalized, and Critical compartments are three additional compartments for further accurate analysis. This model allows the asymptomatic cases, overflowing hospitals with less number of hospital beds and ICU beds. A time-dependent SEAIHCRD model has been divided into eight population-wise compartments: Susceptible (S), Exposed (E), Asymptomatic Infectious (A), Symptomatic Infectious (I), Hospitalized (H), Critical (C), Recovered (R) and Dead (D). A time-dependent SEAIHCRD model describes the transition of people from Susceptible to Exposed, then Exposed to Asymptomatic Infectious and Symptomatic Infectious. Then from Asymptomatic Infectious, they can either Symptomatic Infectious or Recovered compartment. According to the proposed time-dependent SEAIHCRD model, only symptomatic infected individuals can enter the hospitalized compartment and critical compartment. Hospitalized cases may either enter the Critical or Recover compartment and the Critical compartment; they may either die or recover. A time-dependent SEAIHCRD model is shown in Fig. 4.

**Figure 4:**
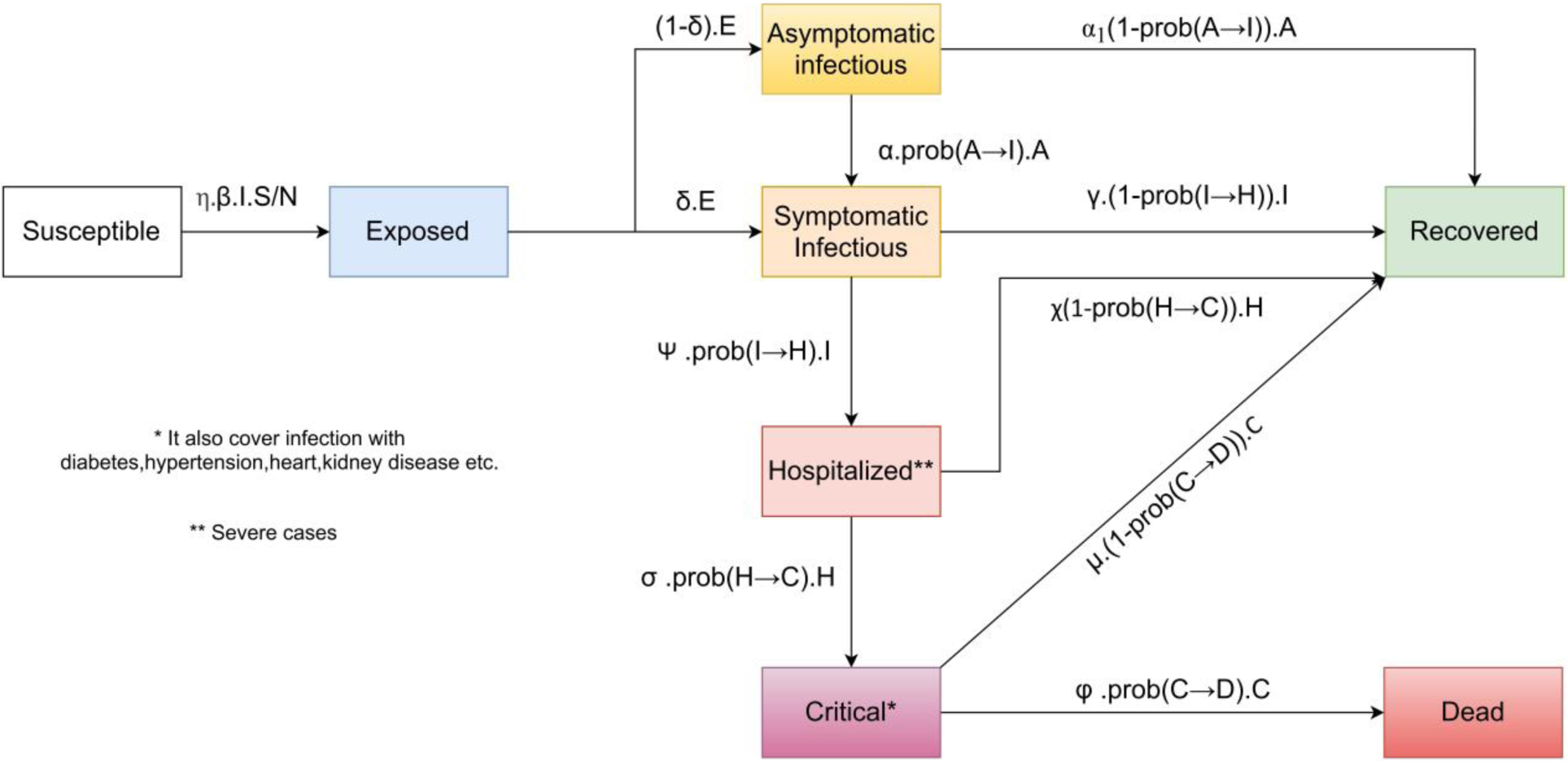
A time-dependent SEAIHCRD model

Asymptomatic cases and some mild cases, put into a home quarantine that does not require hospitalization, these individuals may recover or, if they show some symptoms, go to the symptomatic infectious compartment in the future. People with a serious infection who have severe pneumonia need hospitalization. These people can either recover or move into the critical compartment. People with critical conditions have multi-organs and multiple disorders and need ICU medication. These people either recover or die of the disease.

#### Differential Equations for time-dependent SEAIHCRD model

The SEAIHCRD model’s ordinary differential equations are as follows:

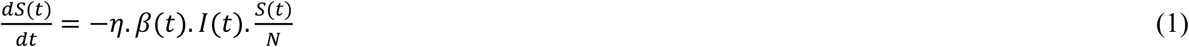

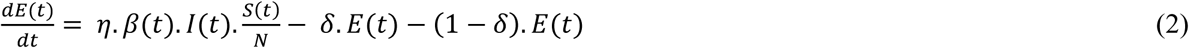

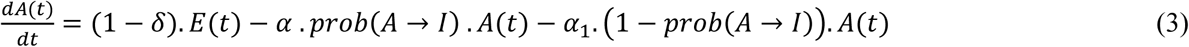

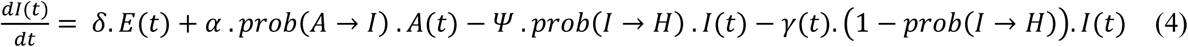

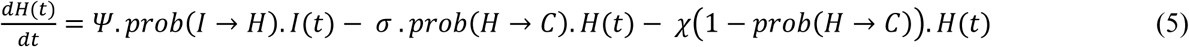

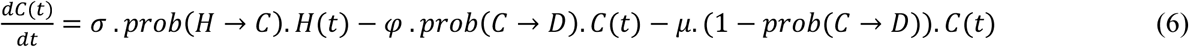

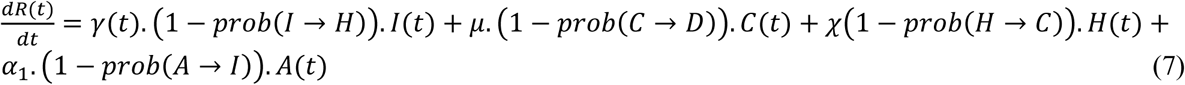

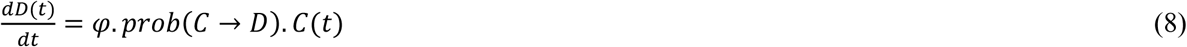

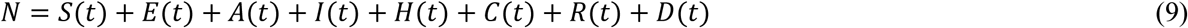

The exact value of the initial conditions (S(0), I(0), R(0)) is not known. We put the amount of the initial conditions very carefully because these systems of ordinary differential equations are extremely sensitive to the initial parameters.

X – Number of days an infected individual has and may spread the disease 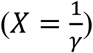

Basic reproduction number *R*_0_ = *β. X*

Therefore, from above, we can write it as 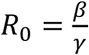

Let β(t) and γ(t) be the transmission rate and recovering rate at time t.

Here

N – Total population

S – A proportion of the entire population that is healthy and has never been infected

E - A proportion of the whole population exposed to infection, transmit the infection and become Symptomatic or purely asymptomatic without being detected

A - A proportion of the population that has no symptoms found positive

I - A proportion of the population with positive symptoms

H - A portion of the whole population, which is positive in the test and hospital

C - A proportion of the population is seriously ill and requires ICU

R - A proportion of the population that has recovered from the infection

D - A proportion of the whole population who died from the infection.

The description of some of the notations used in the time-dependent SEAIHCRD model’s differential equations is:

η – Social distancing factor

β – Rate of transmission

γ-The Recovery Rate

*δ-* The rate of infection transmission from exposed to infectious

*α –* Median time to develop asymptomatic to symptomatic symptoms

*α*_*1*_ *-* Recovery period of asymptomatic cases

*Ψ –* Median time for developing pneumonia and other hospitalization symptoms

*σ –* Median time to ICU admission from the hospital

*φ -* Median Intensive Care Unit (ICU) length of stay

χ - Median hospital stay

*μ -* The time of recovery for critical people

We know that there are limited resources of hospital beds and ICUs available in every country. Sometimes the number of critical cases is more than the number of ICUs. In this condition, doctors have to sort and classify the patients to identify priority and place of treatment. All these conditions are considered in this manuscript.

If a B number of ICUs and a C number of critical cases exist, then the following condition occurs:

1. If there are more ICUs than in severe cases, all patients will be treated.
2. The number of ICUs is less than the number of critical cases (C > B), and the number of patients treated will be B, and the rest (C-B) dies due to shortages.

A time-dependent SEAIHCRD model with critical case analysis is shown in Fig. 5.

**Figure 5:**
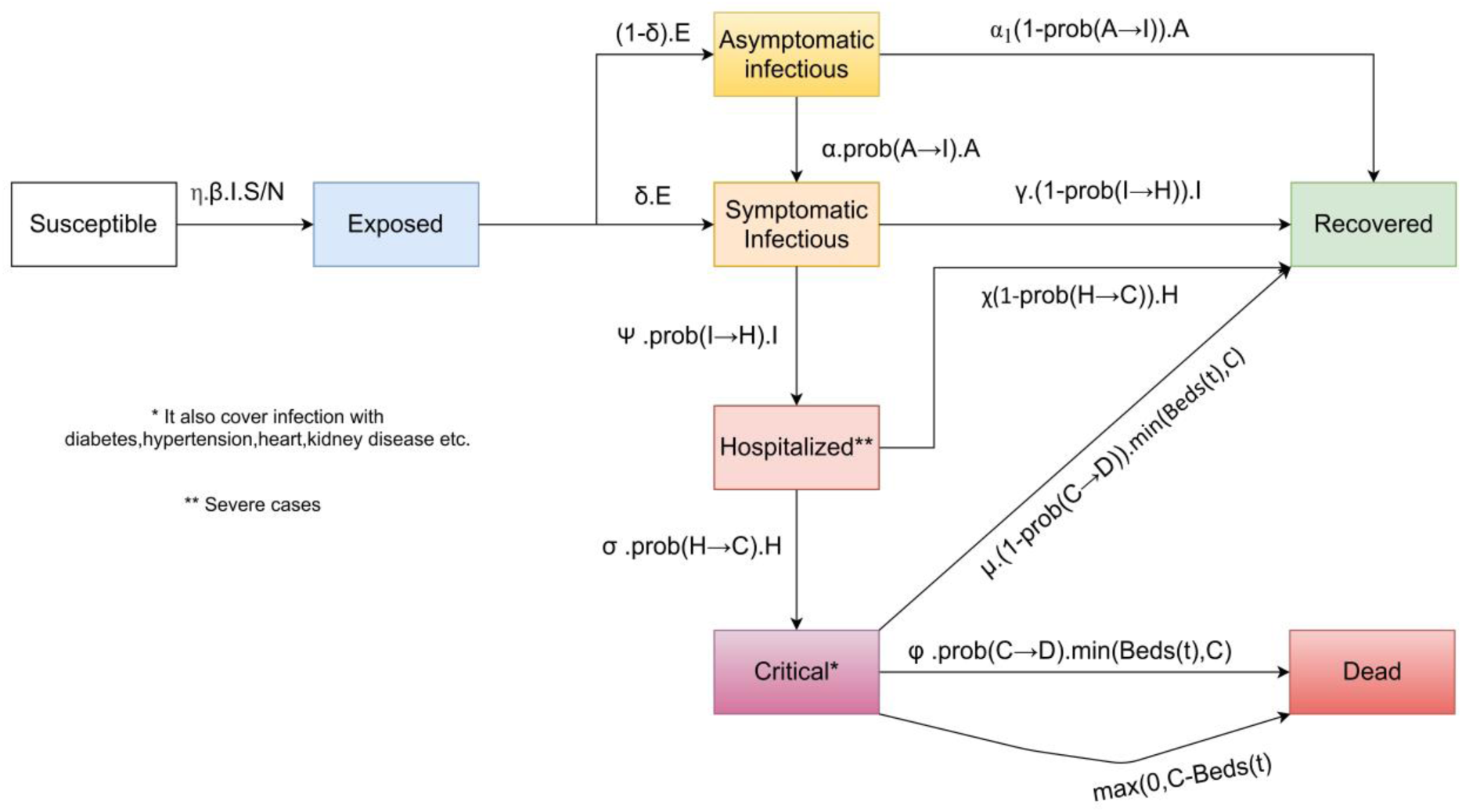
A time-dependent SEAIHCRD model with critical case analysis

The SEAIHCRD model with critical case analysis will modify the ordinary differential equation numbers (6), (7) and (8). Updated equations are as follows:

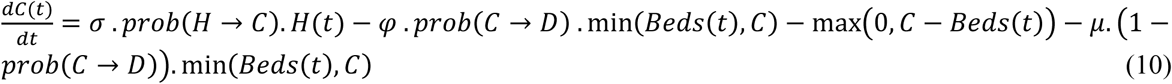

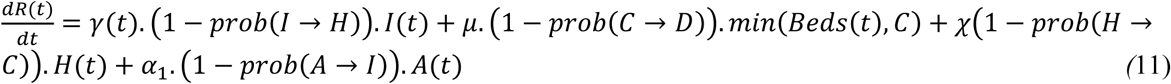

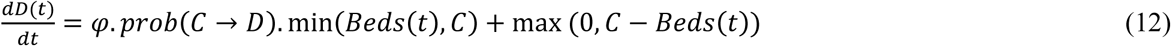

We know that COVID-19 data is updated in days [63]. Hence, we update the differential equations (1), (2), (3), (4), (5), (10), (11), and (12) into discrete-time difference equation:

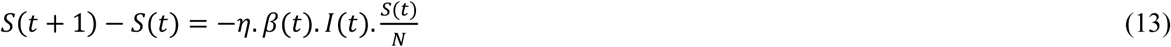

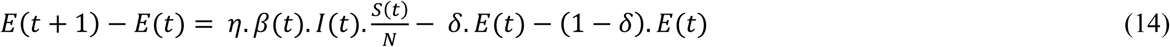

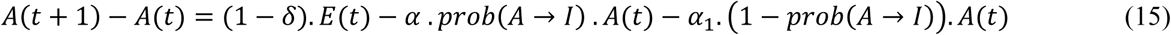

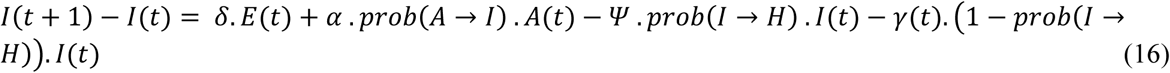

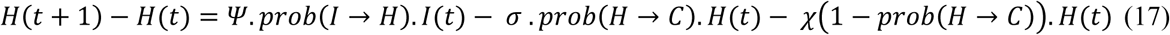

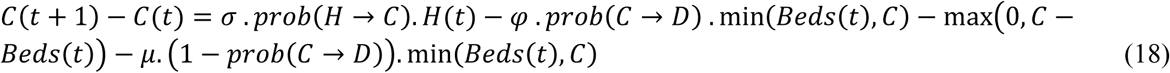

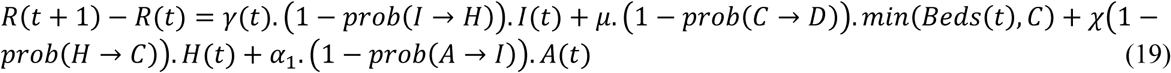

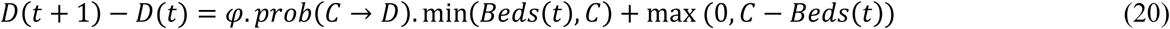

The number of confirmed cases is very low at the very beginning of the spread of the disease, and the majority of the population is susceptible. Hence, for our analysis of COVID-19 in the early stage, we assume that initially, total cases are equal to susceptible cases {S(t) ≈ n, t ≥ 0}.

## 3. Data Analysis and parameter estimation

Compartmental models simplify the mathematical modelling of infectious diseases. Compartmental models are based on ordinary differential equations. A compartmental model can also be used with a stochastic framework that is more accurate but much more complex to analyze. Some initial conditions and characteristic parameters are required to solve these ordinary differential equations. These initial parameters are very sensitive. A small change in them may make a significant difference in outcomes, so they must be carefully introduced in the equations. The distribution of COVID-19 is determined by the nature of several variables in the compartment. Preliminary parameter estimation helps to solve essential consequences like the fatality rate and basic reproduction rate, allowing us to understand more deeply the COVID-19 transmission pattern. In this article, we first collect data for a specific period, then estimate the basic reproduction number, the rate of infection, and the rate of recovery of COVID-19. Based on these estimates, we analyze the spread and endpoint of COVID-19.

The social distance parameter means that the transmission of infectious diseases will avoid social interactions and physical interaction. η is the parameter of social distancing. The social distancing factor value lies between zero and one. When a lock is implemented, and all people are put in quarantine, its value is null, and its value is one if they follow unrestricted and the routine.

### The basic reproduction number R_0_ and case fatality rate over time

The basic reproduction number R_0_ is the ratio between the transmission rate and the recovery rate. The basic reproduction number is the average number of people who can potentially be infected by an infected person if no intervention measures are in place. The value of the basic reproduction number is significant, which is very useful in the intervention of the disease. The value of basic reproduction number R_0_ changes with time; whenever any country adopts lock-downs, the basic reproduction number value decreases, and when that country removes the lock-down, its value begins to increase again. For strict lock-down, the value of R_0_ can be reduced to lower than unity.

#### Transmission Rate β (t) and Recovering Rate γ(t) by Least Square method

In this subsection, we track and predict *β*(*t*) and *γ*(*t*) by the commonly in linear systems. Denote by 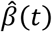 and 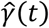 the *predicted* transmission rate and recovering rate. From the FIR filters, they are predicted as follows:

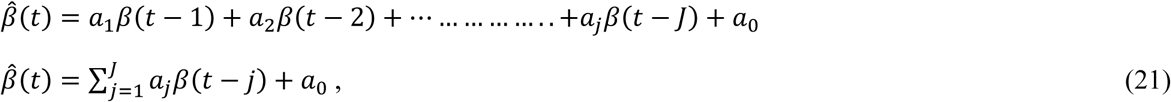

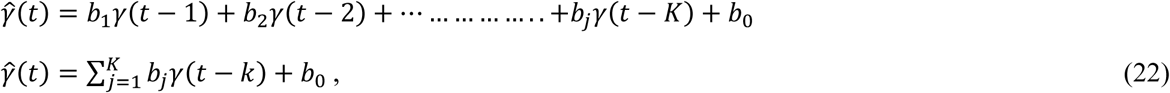

Where *J* and *K* are (0 *< J, K < T −* 2), *a*_*j*_, *j* = 0, 1,*……,J*, and *b*_*k*_, *k* = 0,1,……., *K* are the coefficients. There are several widely used machine learning methods for the estimation of these coefficients, e.g., ordinary least squares, regularized least squares (i.e., lasso regression, ridge regression), and partial least squares [64]. We use the least square as our estimation method, which solves the following optimization problem in the present manuscript.

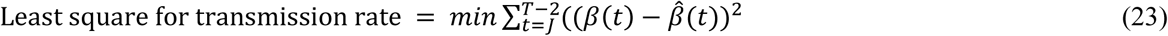

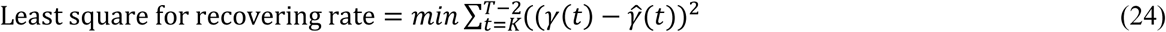

R_0_ is the ratio of β and γ in the compartmental models (R_0_=β/γ). Hence, we can say that *β* = *R*_0_. *γ*. In this manuscript, before the lock-down, the value of R_0_ is R_0_ start, and if the lock-down is imposed on day L, the value of R_0_ is reduced to R_0_ end, and the value of β will be changed accordingly. In real life, the basic reproduction number R_0_ is not constant and changes over time. When social distancing is imposed or removed, its value will change accordingly. The model illustrates the initial impact of social distancing on the basic reproduction number. The time-dependent variation in R_0_ can be given as

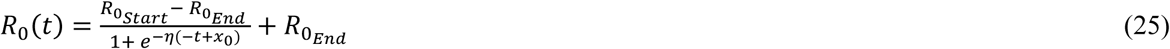

The description of the parameters is given below:

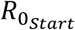 – The value of the R_0_ for the first day

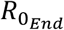 - The value of R_0_ for the last day

x_0_ is the day when R0 drastically decreases or inflection point

η is the parameter of social distance.

The value of the x_0_ should be given very carefully because it makes a big difference to the result and the value of the 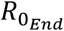 must be less than 1.

We know that (R_0_=β/γ) then

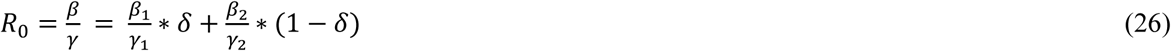

Where β_1_ is the asymptomatic infectious rate, γ_1_ is the asymptomatic recovery rate, β_2_ is the symptomatic infectious rate, and γ_2_ is the symptomatic recovery rate. The combined probability of asymptomatic *δ* and symptomatic infectious disease (1-*δ*) from the exposed compartment is one ((*δ*)+(1-*δ*)=1).

### Age-dependent fatality rate

Infection Fatality Rate is the ratio of the number of deaths to the number of cases. The rate of fatality is not stable and depends on many factors, such as the age factor, fatal diseases. The number of available ICU beds is a common reason, which mostly affects the rate of fatalities. If the majority of people are infected, then the fatality rate is high, and when fewer people are infected, the fatality rate is low. We need more health care if infected people are very high. Sometimes because of the absence of medical facilities, everyone is not treated. Therefore, we need to know what proportion of people are currently infected. Therefore we describe the fatality rate φ as

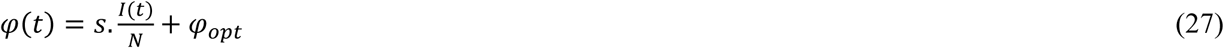

The description of the parameters is given below:

*s* - It is arbitrary, but the fixed value that controls infection

φ_opt_ - optimal fatality rate

The value of parameter *s* is significant as it controls the above equation; very slight changes in s can make a massive difference in results. Therefore, the value of this parameter should be chosen carefully. *I(t)* is the infected at time t, and N is the total population.

Case fatality analysis is complex, depending on the age group. Therefore, we divide age into different age groups to better analyze the case fatality (e.g. people aged 0 to 9, aged 10-19,….., aged 90-100). Two things are required for the age group analysis are the fatality rate by age group and the proportion of the total population in that age group. If the population of older people is higher, then the death rate will be increased, and if the proportion of young people is higher, then the death rate may be low.

Therefore, the Death (fatality) rate is calculated as follows:

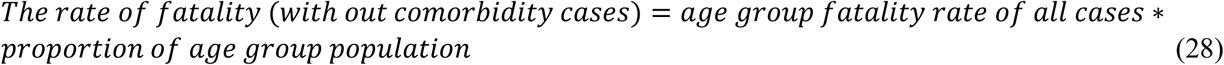

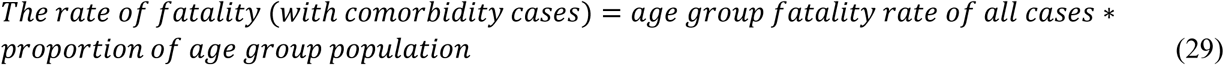

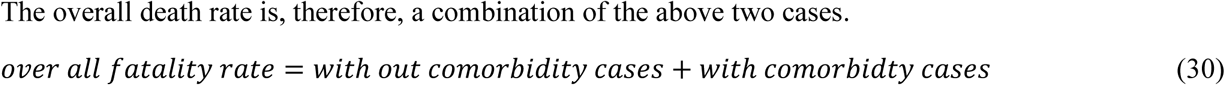

All the cases of comorbidity are considered critical cases in this manuscript. The death rate is the ratio between the number of deaths and the number of cases. The probability of dying may be described as if infected by a virus (in percentage) and depends on various age groups. The following table presents the percentages of deaths if a person is infected with COVID-19 in a given age group [45].

**Table 1:**
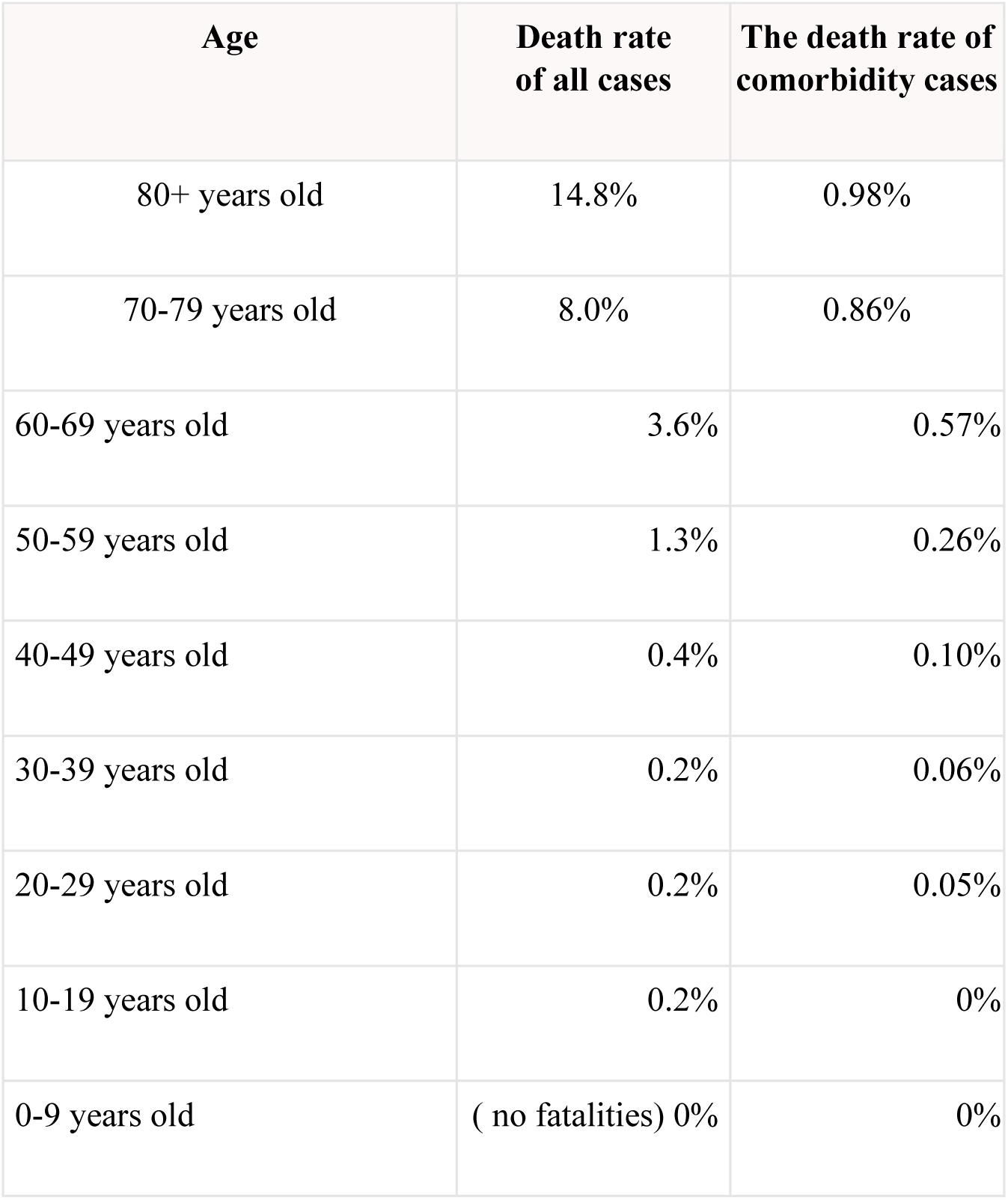
Age-wise death rate and comorbidity in all cases

### Hospitalized and critical cases analysis

We know there is a limited number of hospital beds and ICU beds in every country. All countries are starting new hospitals, opening rooms, etc. to increase the number of hospital beds and ICU beds when the disease spreads. Hence, the number of hospital beds and ICUs is growing over time. We can model the number of beds as follows:

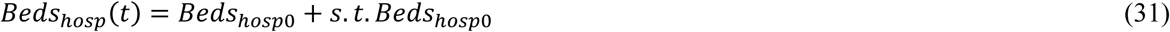

Description of the parameters is given below:

Beds_hosp0_ - Total number of Hospital beds available

*s* - some scaling factor,

and for ICUs

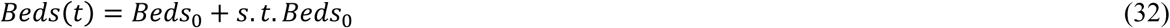

Description of the parameters is given below:

Beds_0_ - Total number of ICU beds available

*s* - some scaling factor

As the number of beds shown in the formula increases times per day to determine how many beds are served for the patients in hospitals each day, this is an important factor.

### Fitting the model to find some important parameters value

In this manuscript, we focus on fitting a time-dependent SEAIHCRD model with time-dependent basic reproduction number and fatality of age group, hospital beds, and ICUs with actual COVID-19 data, which come close to the real data and find parameters for our model that can be useful in the future discussions.

The PYTHON platform used some important libraries such as pandas, NumPy, LMfit, ode, etc. for all the experimental studies. The processor Intel(R) Core (TM) i7-5500U <u>CPU@2.40</u> GHz, 2401 MHz, 2 Core(s), and 4 Logical Processor(s), 12 GB RAM hardware configuration is used for simulation work.

The initial assumptions for the parameters are very crucial. First, we need to know what parameters are known and what we need to get out of it. In this proposed model, many parameters have been used. We will calculate some of the parameters and consider some according to the current study and data. There is no need to fit N, just put the population in the place that we need to model. Likewise, one needs not to calculate beds_0_; simply place the number of ICU beds on the model we have. We value some parameters, i.e. the social distance parameter η, in the range 0 to 1, according to the actual data and analysis, where 0 indicates that everyone is locked and quarantined while one is for a regular lifestyle.

The average incubation period for the virus is 5.2 days but varies between different populations, according to a study by China [46]. For those exposed to infectious agents, the Chinese team study found that 14 days of medical examinations are required. Median hospital stay is around ten days (χ=1/10), the recovery period is around10 days (γ=1/10), the median time to develop asymptomatic to symptomatic symptoms is approximately one week (*α=*1/7), and the recovery period of asymptomatic cases is around ten days (*α*_*1*_*=*1/10). It takes around five days from the appearance of symptoms to the growth of pneumonia [49, 50], and about severe hypoxemia and ICU admission, the median time between the appearance of symptoms is about five to ten days [50-54]. Therefore the median time to develop pneumonia and other symptoms of hospitalization (Ψ=1/5) and hospital to ICU admission (σ=1/7) are used in this study [47-48]. Finally, for COVID-19 patients, the length of stay in intensive care units (ICUs) was approximately 8-13 days, and the critical recovery time was eight days in a Chinese report[54, 55]. Therefore, median Intensive Care Unit (ICU) duration of stay φ =1/8 and median critical recovery time μ=1/8 are chosen.

The case fatality rate (CFR) is the proportion of cases that die of the disease [56]. The basic formula for the case fatality rate is 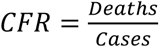 and this formula is sometimes corrected as 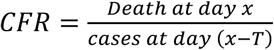, Where T is the average time from case confirmation to death.

We know that *β*(*t*) can be calculated by basic reproduction number *R*_0_(*t*) and *γ*. Hence, no need to find any separate parameter for *β*. The beds scaling factor *s* can be fitted because the number of people being treated due to lack of facility is minimal compared to the number of deaths. Therefore it does not affect much in the result. The proposed model has four estimates of the probabilities *prob*(*I* → *H*), *prob*(*H* → *C*), *prob*(*A* → *I*) and *prob*(*C* → *D*) split by age groups and weighted by age-group proportion. We will try to fit all the above probabilities that are incredibly close to the prediction of a specific risk group, such as diabetics, cancer, heart disease, high blood pressure, etc. now we have to fit all the probabilities *prob*(*I* → *H*), *prob*(*H* → *C*), *prob*(*A* → *I*) and *prob*(*C* → *D*) and 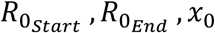 and *η* are the parameters of *R*_0_(*t*).

The Johns Hopkins University Center for Systems Science and Engineering[57] is the principal source of data. We have gathered and cleaned data from UN data[58] for age groups, probabilities, and ICU beds. A total number of Hospital beds and ICUs per 100,000 peoples for topmost affected countries. The number of people per age group for top affected countries. Probabilities for *prob*(*I* → *H*), *prob*(*H* → *C*), *prob*(*A* → *I*) and *prob*(*C* → *D*) per age groups, but we use fitted probabilities and collect data on the number of fatalities on a day-to-day basis from January 22, 2020. As the number of reported cases depends on many factors, such as the number of test cases, etc. and hence usually is not very accurate. However, the reported deaths are more accurate. Therefore, we use fatality statistics instead of total or active cases.

Firstly, we have collected all the related available data and parameters available to us from reliable sources and then defined initial guesses and limits for the remaining parameters of this model within certain limits depending on the situation. We have adopted the Levenberg – Marquard algorithm to extract various parameters from the data fitting as this is the most important and effective reported data-fitting algorithm [59, 60]. For Python, a high-quality interface is provided by the LMfit library for non-linear optimization and curve-fitting problems [61]. The R-squared value measures the proximity of the data to the fitted line. The R-square value is always between zero and one. In general, higher the R-squared value, better the model a given your data. The curve of death cases in Brazil, India, Mexico, Russia, South Africa, and the United States is shown in Fig. 6

**Figure 6:**
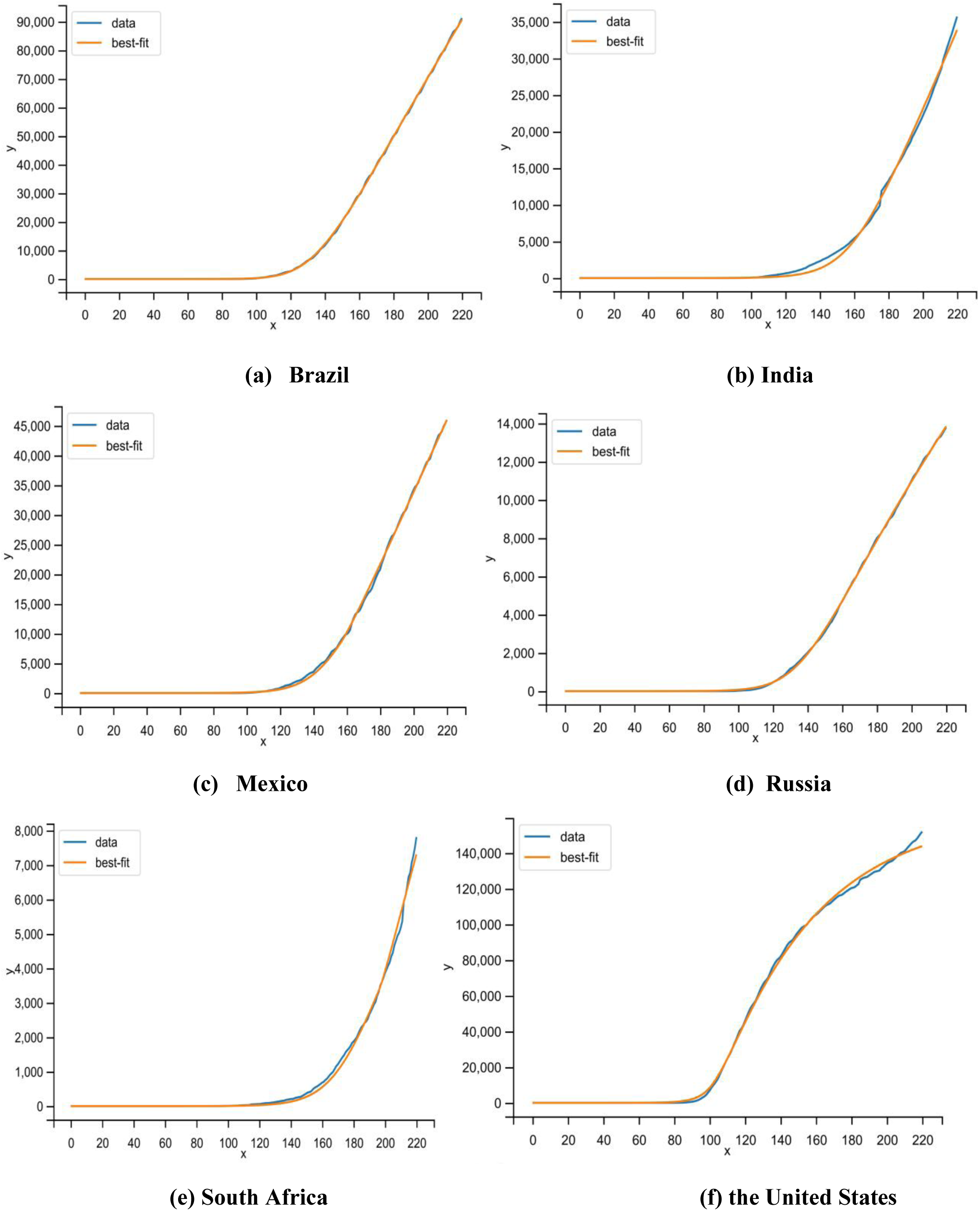
Curve fitting of death cases in Brazil, India, Mexico, Russia, South Africa, and the United States

**Table 2:** R-squared value of curve fitting for Brazil, India, Mexico, Russia, South Africa, and the United States

| Country | R-squared value (Death cases) |
| --- | --- |
| **Brazil** | **.992** |
| **India** | **.962** |
| **Mexico** | **.985** |
| **Russia** | **.988** |
| **South Africa** | **.979** |
| **United States** | **.968** |

According to the study, most cases are mild or asymptomatic and can recover at home with proper guidance. Fewer percentages of people are critical, but if the patient is old or has some pre-existing illnesses such as chronic respiratory disease, diabetes, cancer, hypertension, and cardiovascular disease, the risk is very high. We gather and analyze data provided by New York City Health as of June 1, 2020 [61, 62] are shown below:

**Table 3:** Age-wise rate of hospitalization from infected cases, critical cases from infected cases, and death cases from ICUs

| AGE | Hospitalization rate from Infected cases | Rate of critical cases from Infected cases | Rate of dead cases from ICUs |
| --- | --- | --- | --- |
| 80+ years old | 18.4% | 0.16% | 0.98% |
| 70-79 years old | 16.6% | 0.10% | 0.86% |
| 60-69 years old | 11.8% | 0.07% | 0.57% |
| 50-59 years old | 8.2% | 0.05% | 0.26% |
| 40-49 years old | 4.3% | 0.03% | 0.10% |
| 30-39 years old | 3.4% | 0.025% | 0.06% |
| 20-29 years old | 1.0% | 0.009% | 0.05% |
| 10-19 years old | 0.1% | 0.003% | 0% |
| 0-9 years old | 0.01% | 0.001% | 0% |

We used the publicly available dataset of COVID-19 provided by Johns Hopkins University in this study [83]. This dataset includes the daily count of confirmed cases, recovered cases, and deaths in many countries. The time-series data are available from January 22, 2020, onwards. We also collected and crosschecked data in Worldometer, Coronavirus cases [82], a website providing real-time data for COVID-19. These data are gathered through announcements by public health authorities and report public and unidentified patient data immediately; hence ethical authorization is not necessary.

### Distribution of Total cases worldwide

According to WHO, 216 countries are affected by COVID-19. Fig. 7 shows that 61% of cases are from six countries only. The United States of America has the highest number of cases, which alone is 27% of the world.

**Figure 7:**
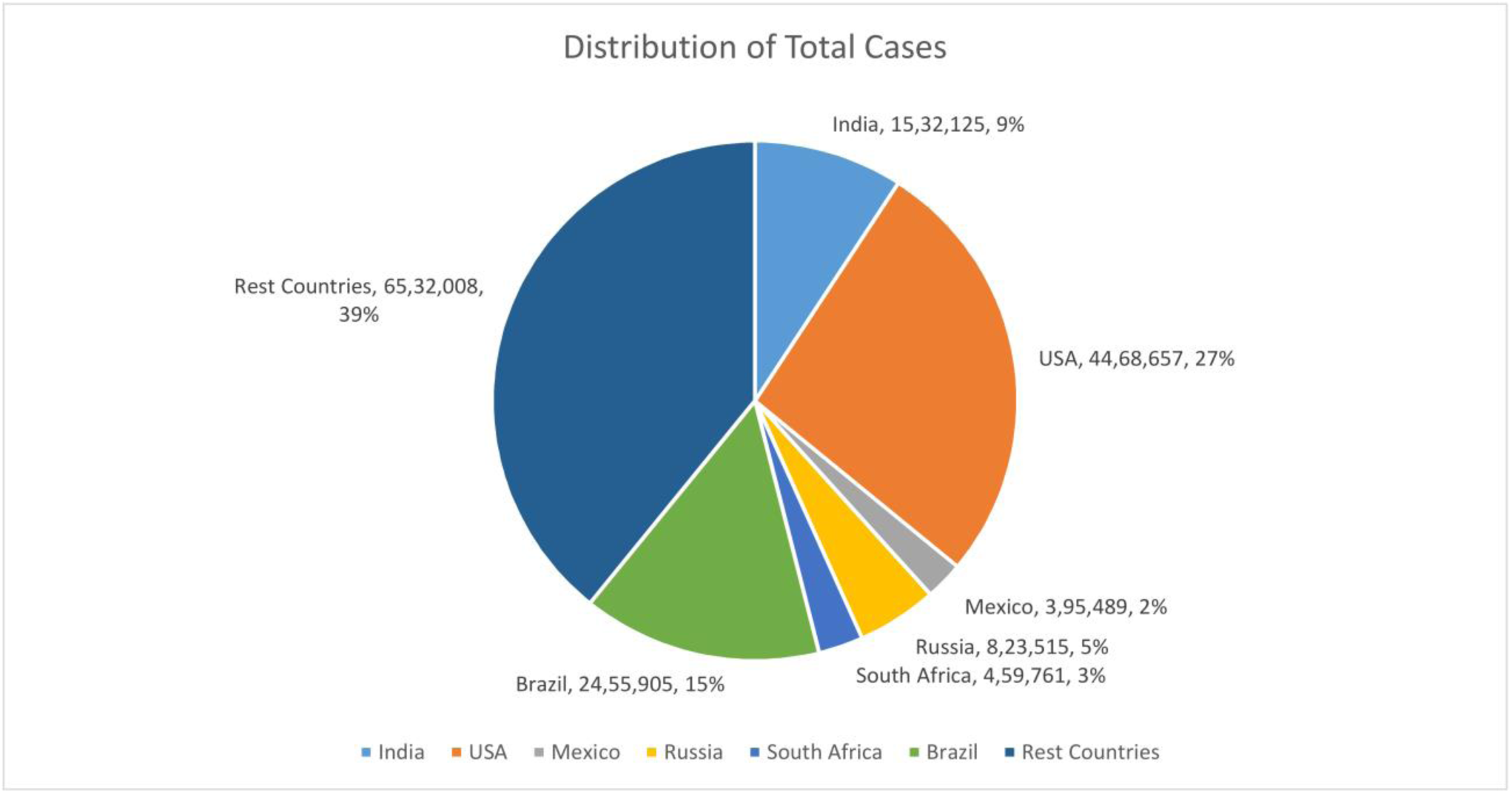
Distribution of total cases worldwide

### Distribution of Death cases worldwide

As discussed above, Death cases are more accurate as there is rarely any chance that death due to COVID-19 is not reported or recorded by the related authorities. Fig. 8 shows that 51% of cases are from six countries only. The United States of America has the highest number of cases, which alone is 23% of the world.

**Figure 8:**
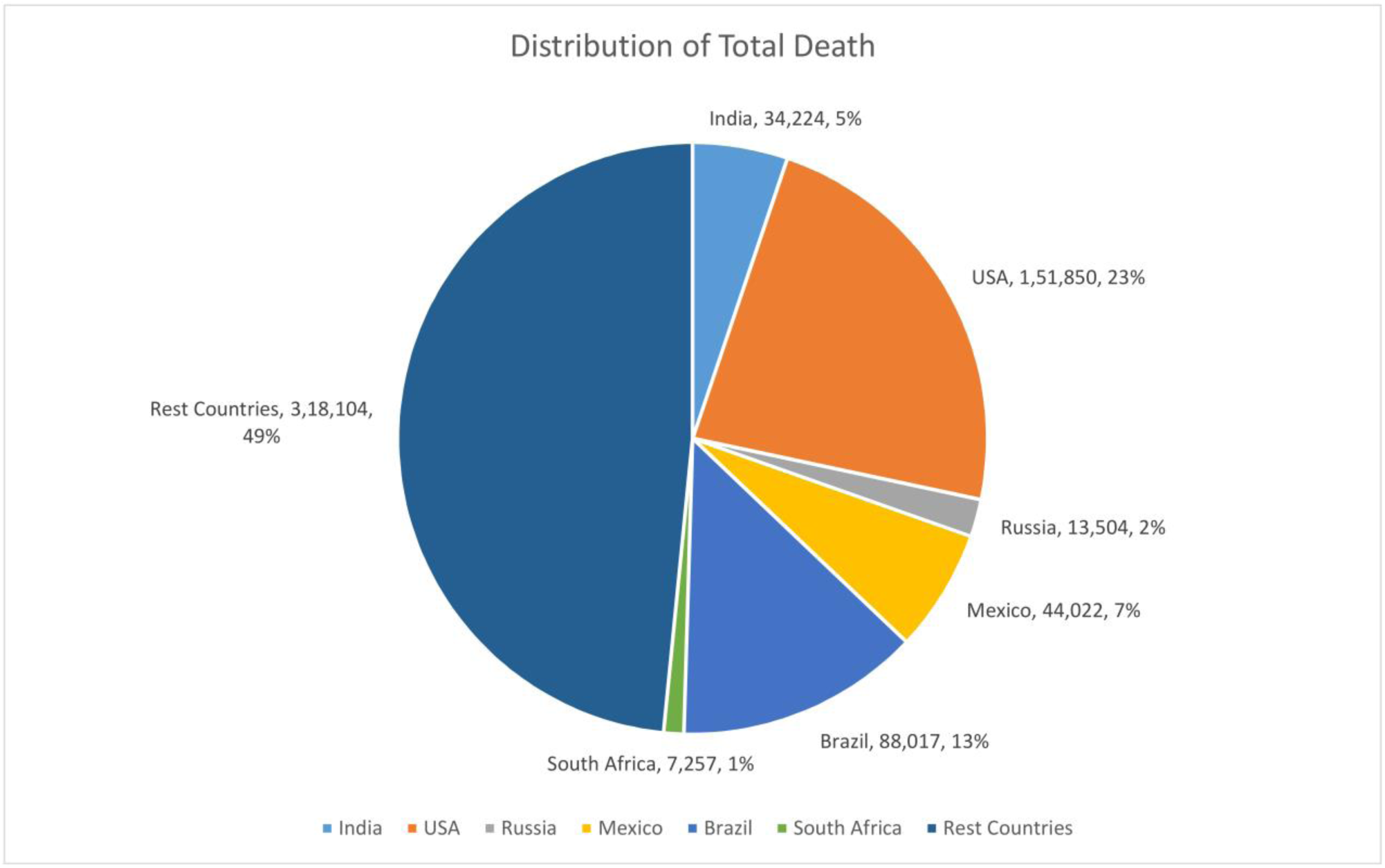
Distribution of Death cases worldwide

### Total cases

The fig. 9 shows the country-wise total cases from January 22, 2020 to July 28, 2020 of COVID-19.

**Figure 9:**
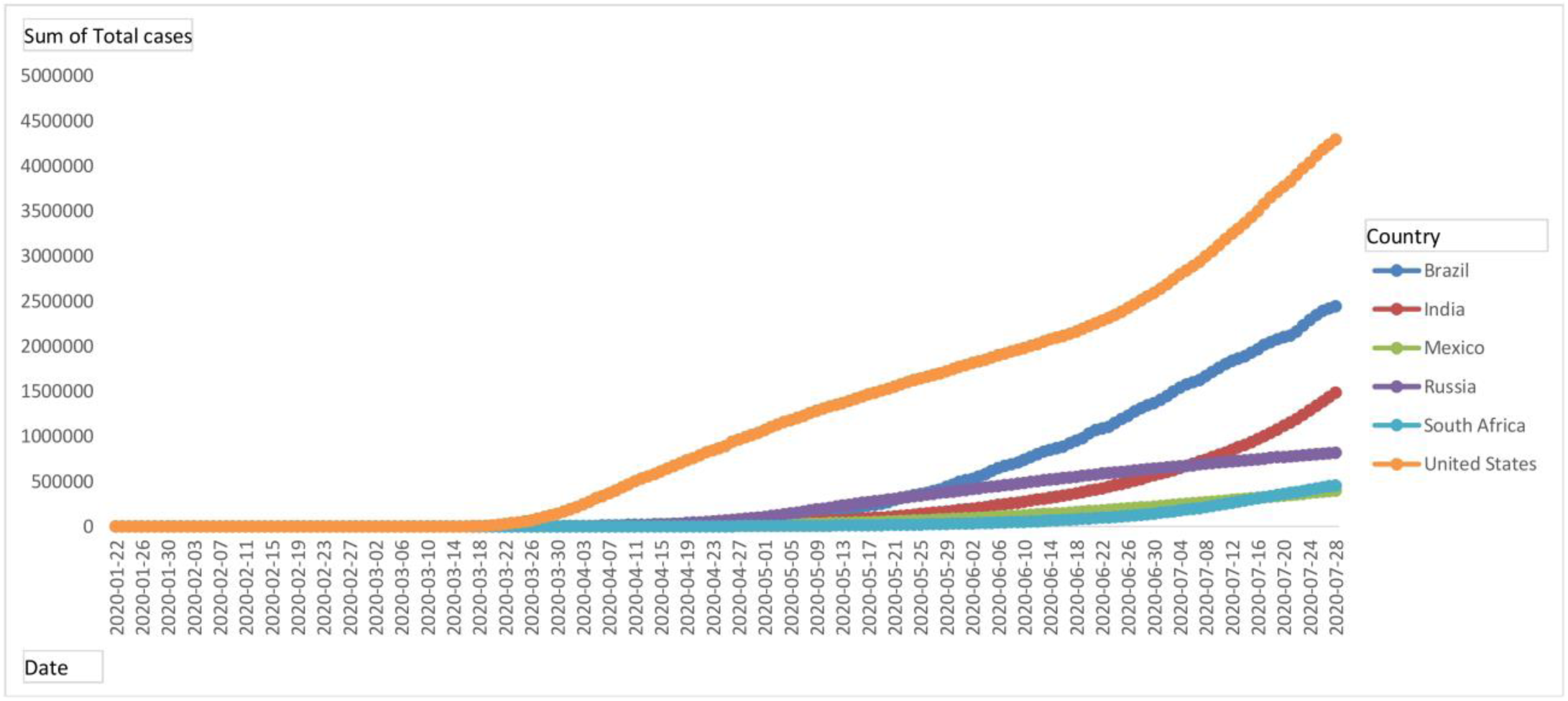
Day-wise total cases of Brazil, India, Mexico, Russia, South Africa, and the United States

### Total death

Fig. 10 shows the country-wise total death from January 22, 2020 to July 28, 2020 of COVID-19.

**Figure 10:**
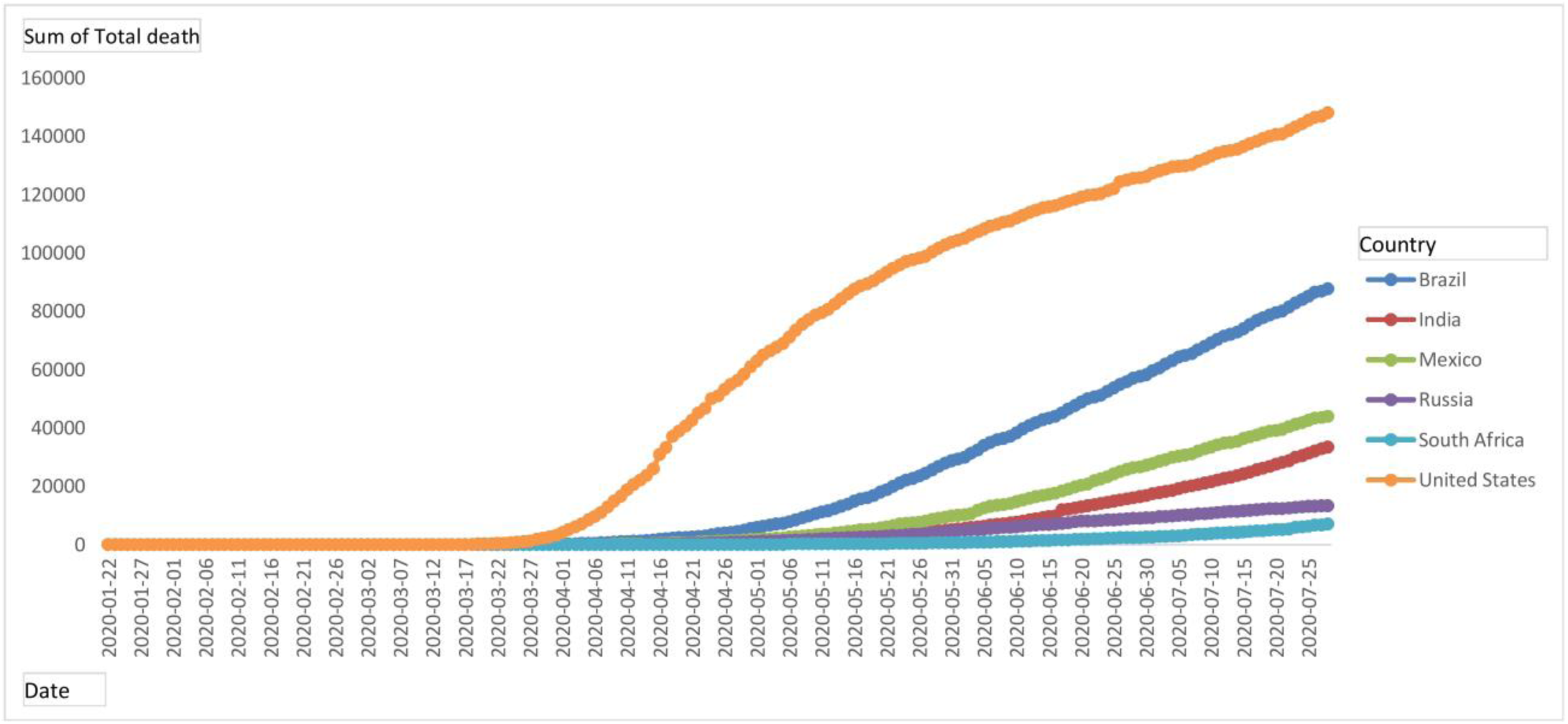
Day-wise total death of Brazil, India, Mexico, Russia, South Africa, and the United States

### Daily new cases

Fig. 11 shows the country-wise daily new cases from January 22, 2020 to July 28, 2020 of COVID-19.

**Figure 11:**
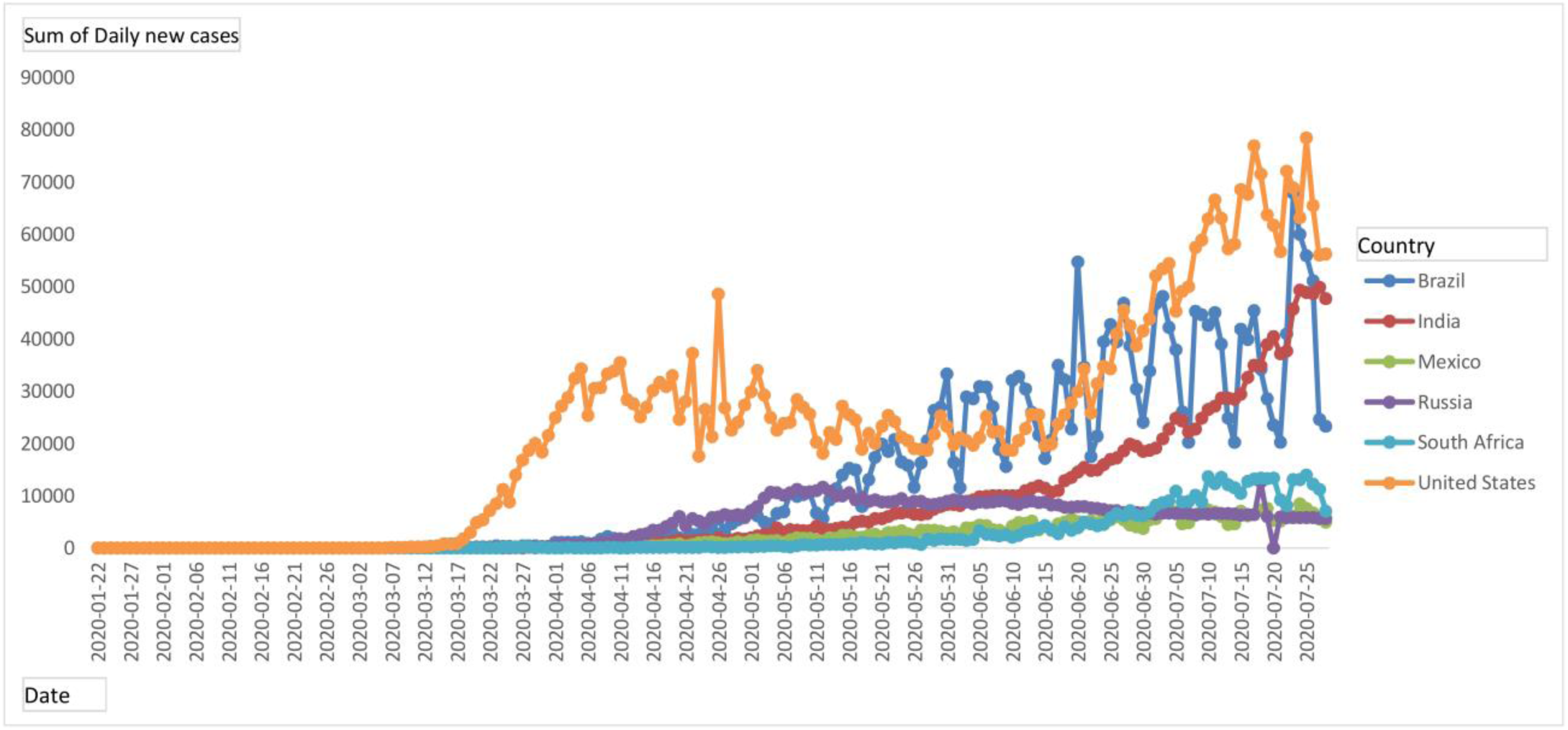
Daily new cases of Brazil, India, Mexico, Russia, South Africa, and the United States

### Daily new death

Fig. 12 shows the country-wise daily new death cases from January 22, 2020 to July 28, 2020 of COVID-19.

**Figure 12:**
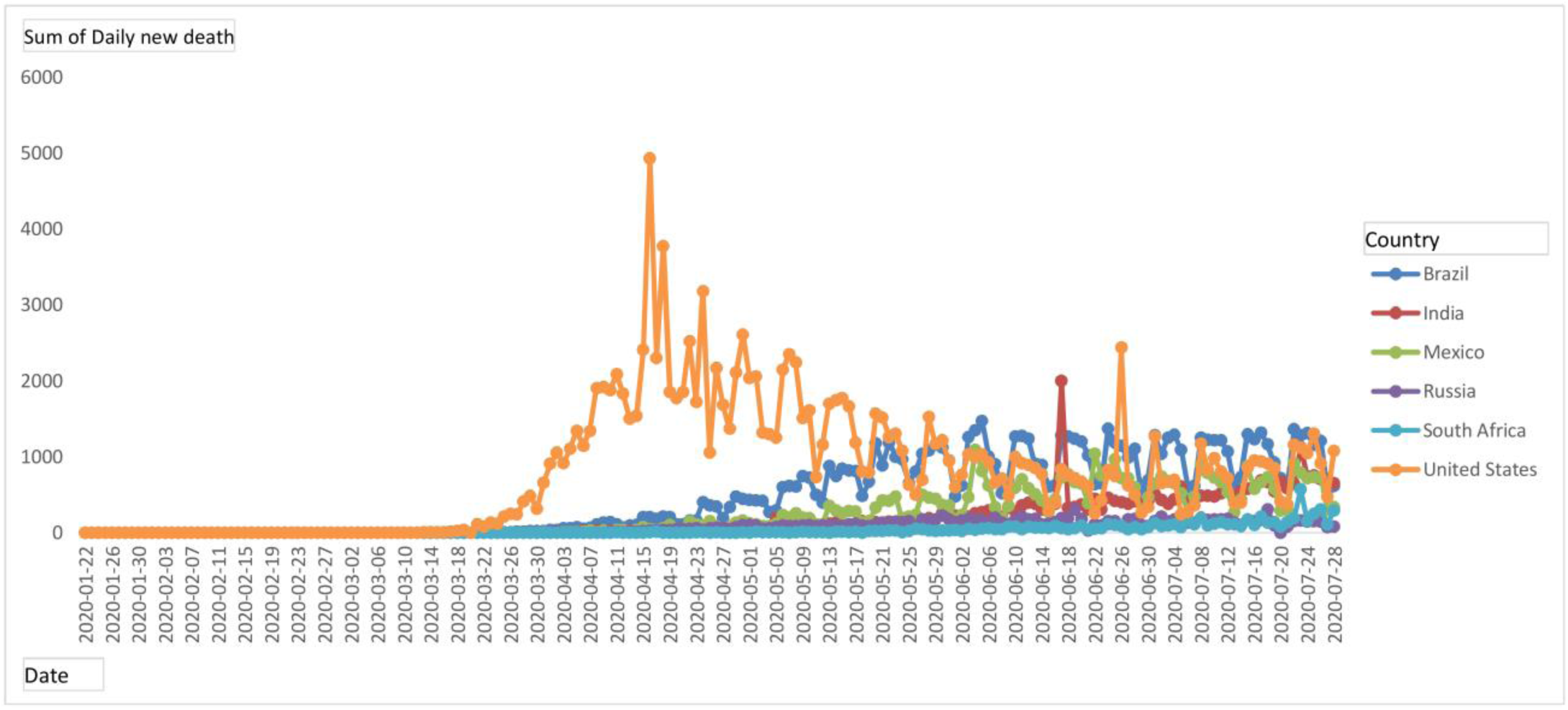
Daily new death cases of Brazil, India, Mexico, Russia, South Africa, and the United States

### Case fatality rate

The overall fatality rate of an ongoing pandemic is difficult to estimate, but the current fatality rate can be calculated. Fig. 13 shows the Country wise case fatality rate as of the current date of July 28, 2020.

**Figure 13:**
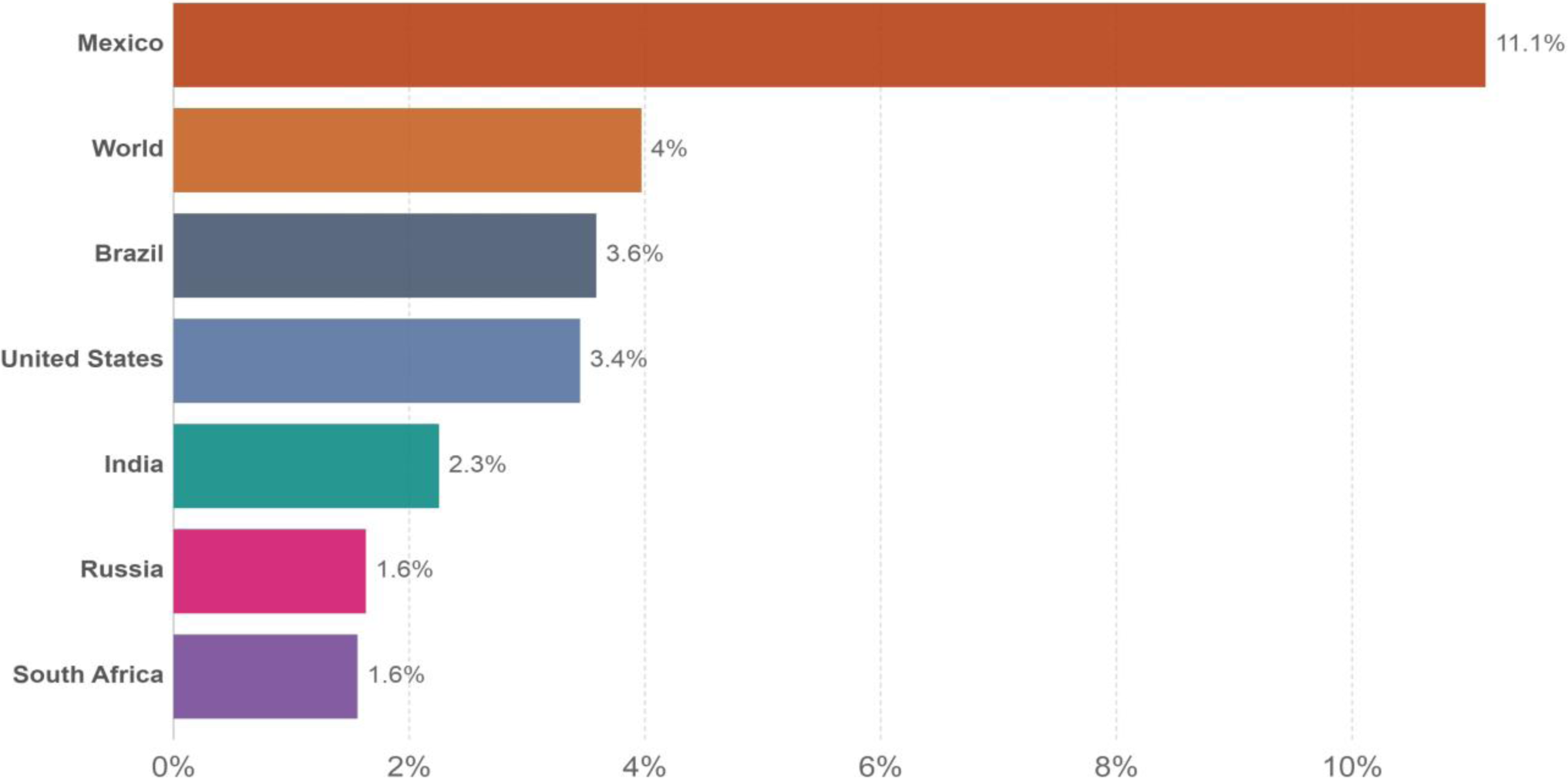
Case fatality rate as of July 28, 2020, for Brazil, India, Mexico, Russia, South Africa, and the United States

### Age-wise population

In terms of the age group, the population of Russia and the United States in 70 years and older is more percentage-sized, according to World Bank data. In the age range of 0 to 20 years, the population of South Africa, India, and Brazil is higher. In all these countries, the population distribution of Brazil, India, Mexico, Russia, South Africa, and the United States is nearly equivalent between 20 and 70 years. The population distribution by age group is represented in Fig. 14.

**Figure 14:**
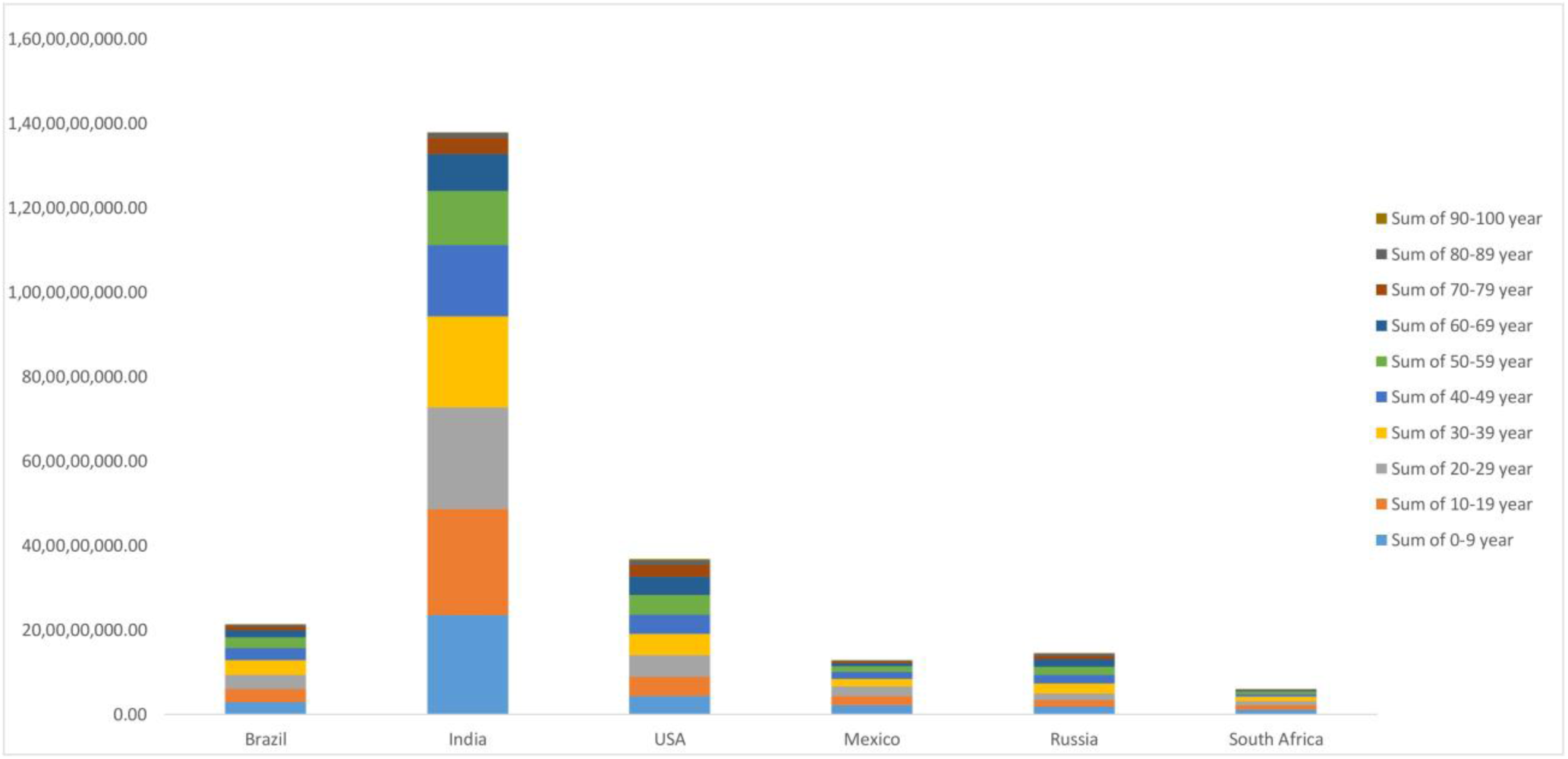
The distribution of the population according to the age category of Brazil, India, Mexico, Russia, South Africa, and the United States

### The number of hospital beds per 100k

Unit beds per 100,000 population or a 100k population, according to WHO and the OECD. Basic measures focus on all hospital beds that are occupied or empty. The Country-wise number of hospital beds is shown in Fig. 15.

**Figure 15:**
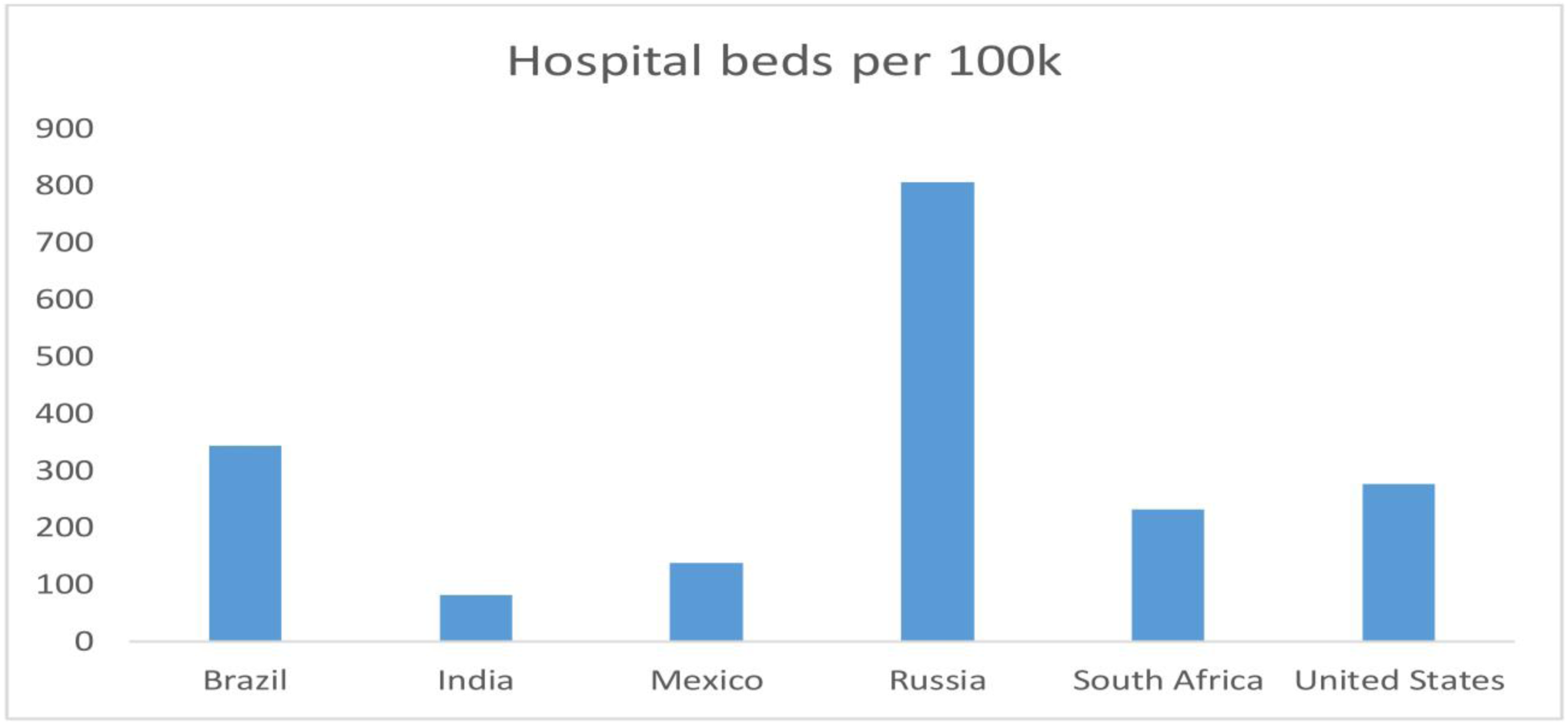
The Country-wise number of hospital beds of Brazil, India, Mexico, Russia, South Africa, and the United States

### The number of ICU beds per 100k

ICU Bed Counts are made up of beds per 100,000 population or 100,000 population, according to WHO and OECD. ICU beds are provided for all cases with severe conditions. The Country-wise number of ICU beds is shown in Fig. 16

**Figure 16:**
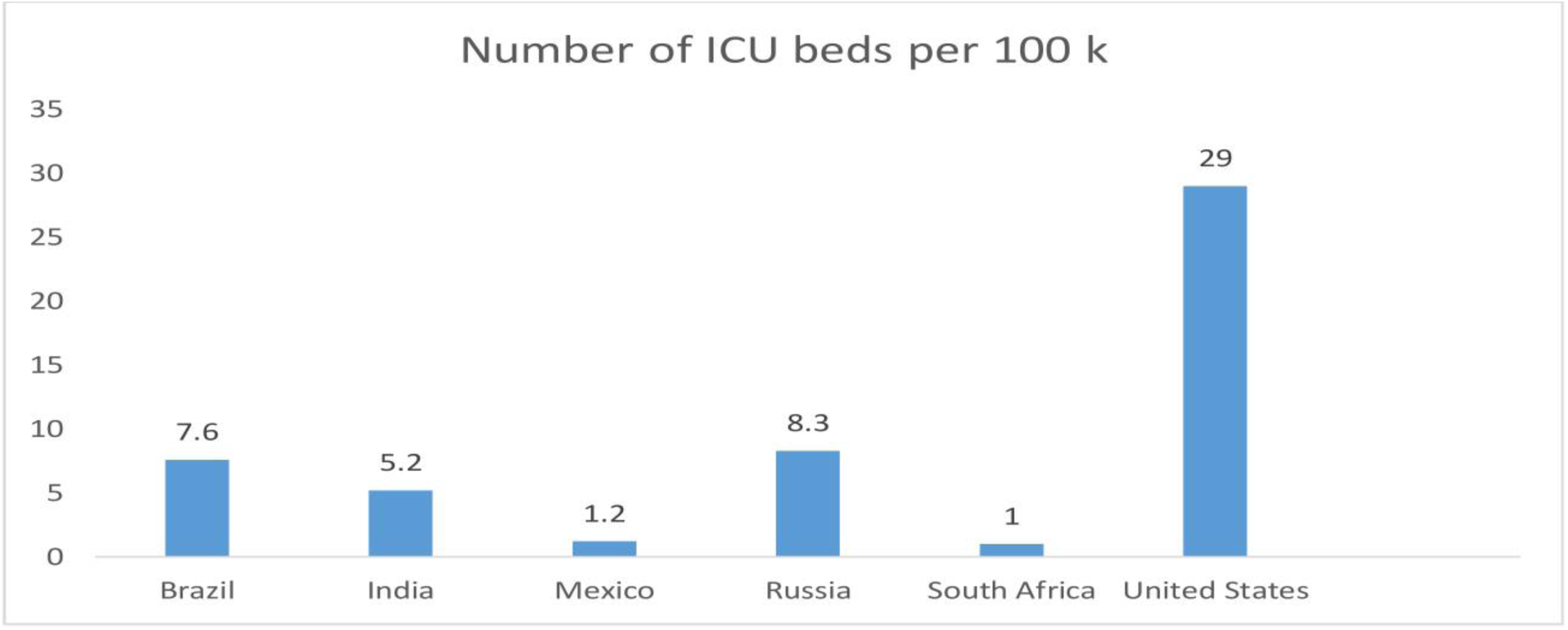
The Country-wise number of ICU beds of Brazil, India, Mexico, Russia, South Africa, and the United States

## 4. Results and Discussion

We analyze and forecast in this section, using our proposed A time-dependent SEAIHCRD model, the pattern of COVID-19 among the world’s most affected countries. The result of our proposed method shows that Brazil, Russia, South Africa, and the United States have experienced their worst time. India and Mexico have yet to see a peak at the end of August. If the number of cases is very high in the middle and late August, hospital beds and ICU beds will be needed in India and Mexico. Geographical conditions are entirely different in some countries. The distribution of cases in these areas is also very random. In some places, it spreads too much, and in some places, it spreads far less. In India, the distribution of disease is very random, e.g., the number of cases in Maharashtra, Tamil Nadu, Andhra Pradesh, Karnataka, and Delhi is very high as compared to other states. According to the proposed method, cases in India will start to decline continuously in the first week of September, and the cases will end completely in the first week of November. If we compare the results of our proposed method with real-world data, it is mostly the same. In this section, we calculate for each country the basic reproduction number, the case fatality rate, the number of hospital beds required at peak time, and the number of ICU beds required when serious cases are high.

### 4.1 A time-dependent SEAIHCRD model for Brazil

Brazil is one of the most affected countries by the COVID-19 disease, just behind the United States as of the starting of June 2020. According to the proposed method, cases in Brazil will start to decline continuously in the first week of August, and the cases will end completely in the first week of November. The cases reach a peak in Brazil, according to our proposed method, in July and August. The result of our proposed method shows that Brazil will need some Hospital beds and ICU beds in June, July, and August, when the number of cases is at a peak. The result shows the basic reproduction rate is 4.0 in starting after some time, it starts decreasing visually and goes up to one and even lower. A time-dependent SEAIHCRD model for Brazil shows that the case fatality rate of COVID-19 in Brazil is moderate. There are many ups and downs in the daily case fatality rate, but it goes higher and higher up to 5.5 in our model and then slows down, and total CFR is nearly 3.5 overall. The proposed model for Brazil showing that around 200^th^ day, the number of hospital beds and the number of ICU beds are required. That time the number of death cases is 1800 per day, and daily-hospitalized cases are nearly 3100. The proposed method requires approximately 550 hospital beds and 500 ICU beds when the cases are at their peak in Brazil. A time-dependent SEAIHCRD model results for Brazil has shown in Fig. 17

**Figure 17:**
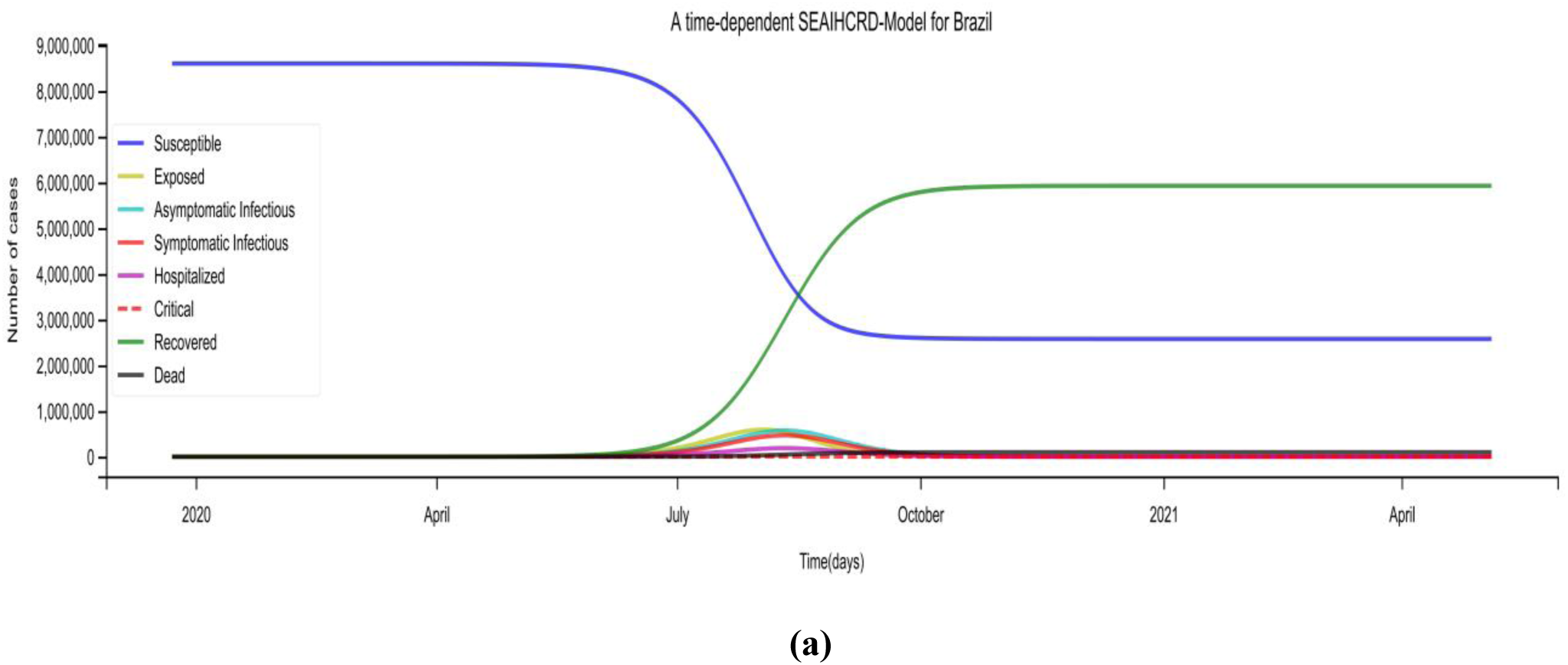
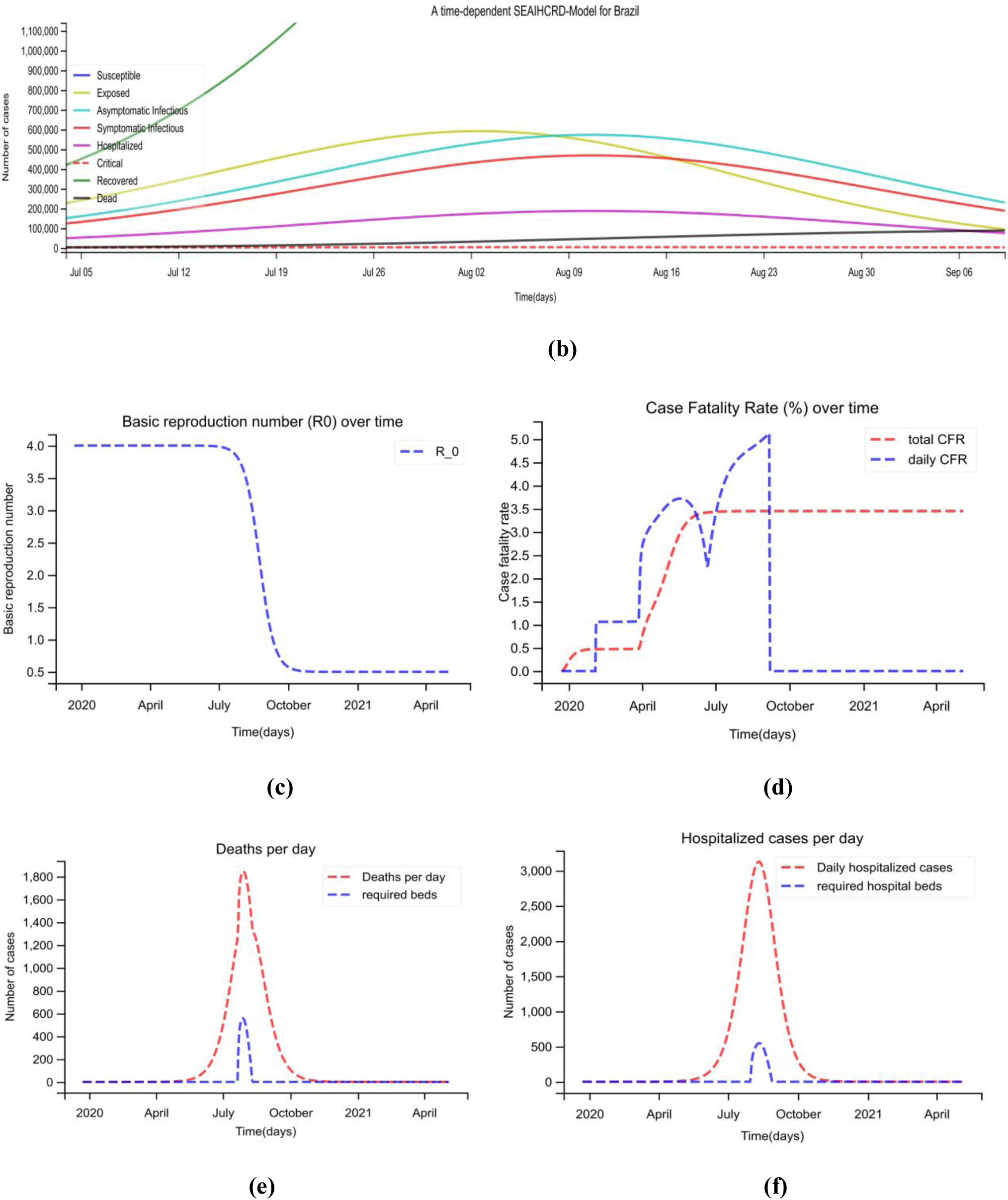
A time-dependent SEAIHCRD model for Brazil. **(a)** The spread scenario of A time-dependent SEAIHCRD model for Brazil. A 500-day analysis has been done by the proposed model, which starts from January 22. **(b)** In this, the peak point of the model has shown by zooming for cases. **(c)** The basic reproduction number of Brazil over time **(d)** the case fatality rate of Brazil over time. **(e)** Daily death cases in Brazil, where the red line shows the number of deaths per day and the blue line shows how many ICU beds are required during peak days. **(f)** Brazil’s Daily Hospital Cases, where the red line indicates the number of hospital cases per day, and the blue line shows how many hospital beds are required during peak days.

### 4.2 A time-dependent SEAIHCRD model for India

India is one of the most affected countries by the COVID-19 disease, just behind the United States and Brazil as of May 2020. According to the proposed method, cases in India will start to decline continuously in the first week of September, and the cases will end completely in the first week of December. The cases reach a peak in India, according to our proposed method in August. The result of our proposed method shows that India will need some Hospital beds and ICU beds in July, August, and September, when the number of cases is at a peak. The result shows the basic reproduction rate is 2.2 in starting after some time, it starts decreasing visually and goes up to one and even lower. A time-dependent SEAIHCRD model for India shows that the case fatality rate of COVID-19 in India is meagre. There are many ups and downs in the daily case fatality rate, but it goes higher and higher up to 2.7 in our model and then slows down, and total CFR is nearly 2.2 overall. The proposed model for India shows that around 230^th^ day, maximum number of hospital beds and the number of ICU beds are required. At that time, the number of death cases is 1200 per day, and daily-hospitalized cases are nearly 2600. As per the proposed method needs approximately 700 hospital beds, and 300 ICU beds will be required during the peak time of COVID-19 in India. A time-dependent SEAIHCRD model results for India has shown in Fig. 18.

**Figure 18:**
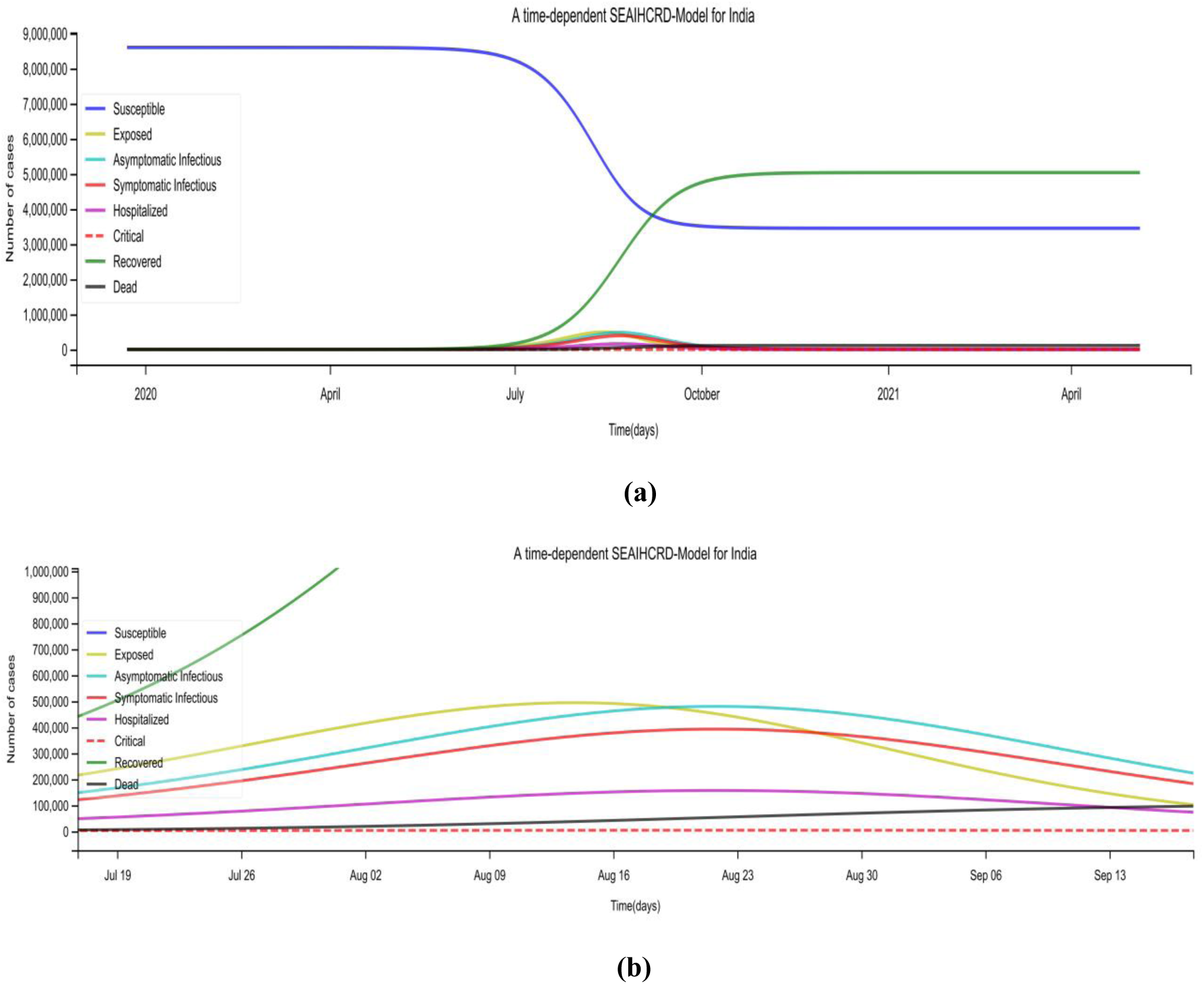
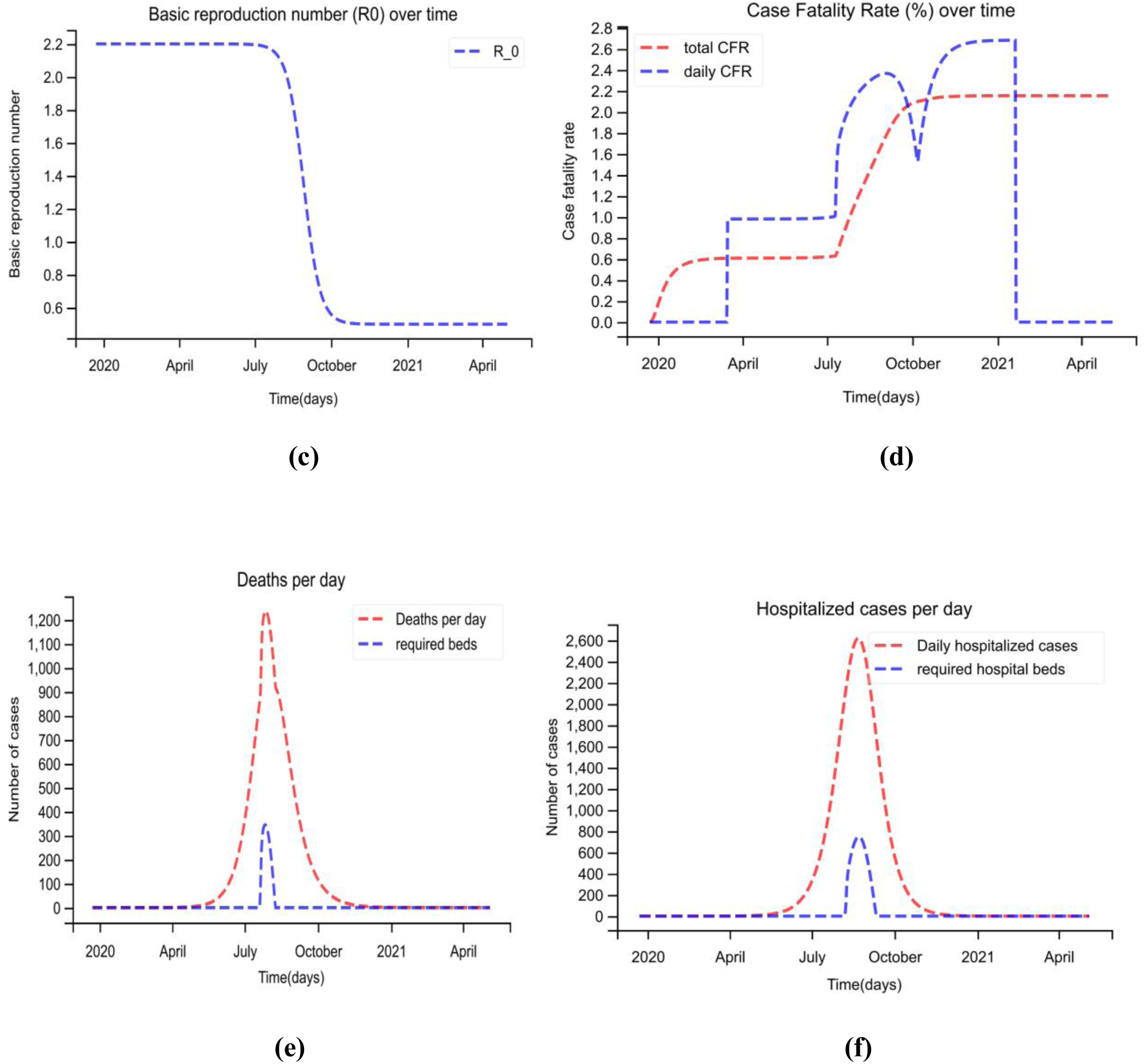
A time-dependent SEAIHCRD model for India. **(a)** The spread scenario of A time-dependent SEAIHCRD model for India. A 500-day analysis has been done by the proposed model, which starts from January 22. **(b)** In this, the peak point of the model has shown by zooming for cases. **(c)** The basic reproduction number of India over time **(d)** the case fatality rate of India over time. **(e)** Daily death cases in India, where the red line shows the number of deaths per day and the blue line shows how many ICU beds are required during peak days. **(f)** India’s Daily Hospital Cases, where the red line indicates the number of hospital cases per day, and the blue line shows how many hospital beds are required during peak days.

### 4.3 A time-dependent SEAIHCRD model for Mexico

Mexico is the fourth most affected countries by COVID-19 disease. According to the proposed method, cases in Mexico will start to decline continuously in the first week of September, and the cases will end completely in the first week of December. The cases reach a peak in Mexico, according to our proposed method, in August and September. The result of our proposed method shows that Mexico will need some Hospital beds and ICU beds in July, August, and September, when the number of cases is at a peak. The result shows the basic reproduction rate is 2.2 in starting, and then after some time, it starts decreasing visually and goes up to one and even lower. A time-dependent SEAIHCRD model for Mexico shows that the case fatality rate of COVID-19 in Mexico is very high. There are many ups and downs in the daily case fatality rate, but it goes higher and higher up to 11.5 in our model and then slows down, and total CFR is nearly 5.0. The proposed model for Mexico showing that around 240th day, the number of hospital beds and the number of ICU beds are required. That time the number of death cases is 950 per day, and daily-hospitalized cases are nearly 650. The proposed method requires approximately 150 hospital beds and 200 ICU beds when the cases are at their peak in Mexico. Mexico’s case fatality rate is very high, so the death cases are very high here, and with more critical cases, more ICU beds will be needed. A time-dependent SEAIHCRD model results for Mexico has shown in Fig. 19.

**Figure 19:**
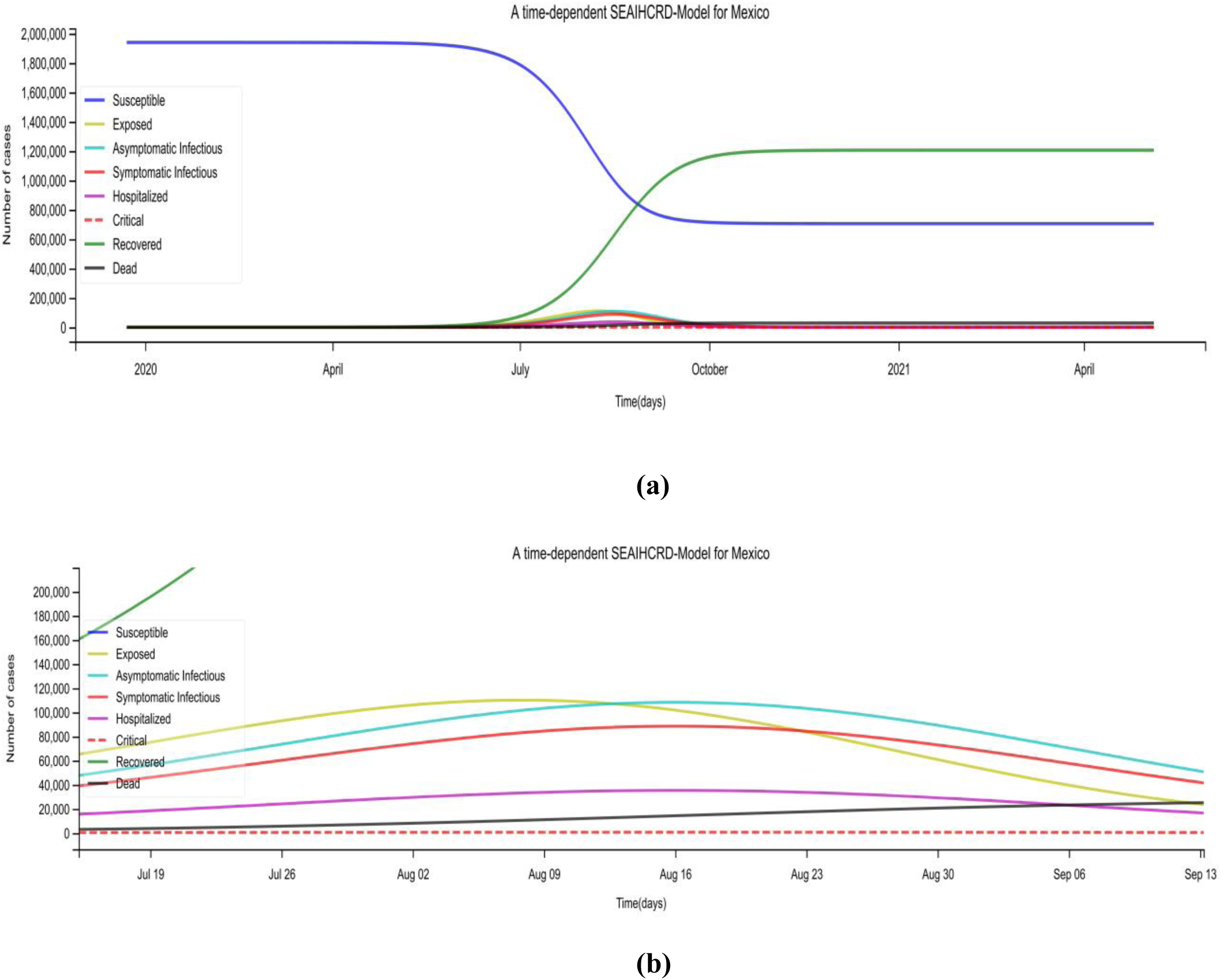
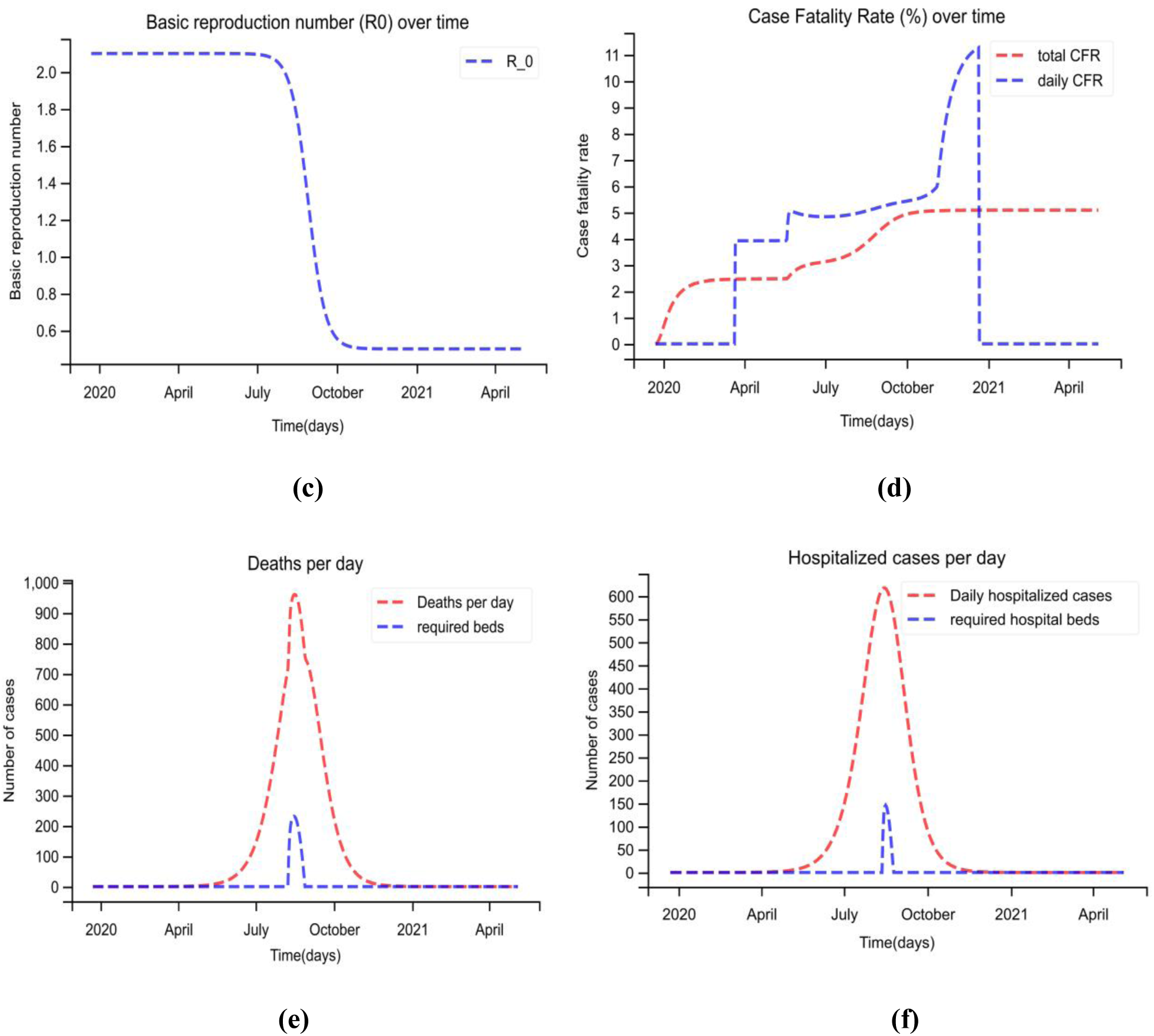
A time-dependent SEAIHCRD model for Mexico. **(a)** The spread scenario of A time-dependent SEAIHCRD model for Mexico. A 500-day analysis has been done by the proposed model, which starts from January 22. **(b)** In this, the peak point of the model has shown by zooming for cases. **(c)** The basic reproduction number of Mexico over time **(d)** the case fatality rate of Mexico over time. **(e)** Daily death cases in Mexico, where the red line shows the number of deaths per day and the blue line shows how many ICU beds are required during peak days. **(f)** Mexico’s Daily Hospital Cases, where the red line indicates the number of hospital cases per day, and the blue line shows how many hospital beds are required during peak days.

### 4.4 A time-dependent SEAIHCRD model for Russia

Russia is also one of the most affected countries by COVID-19 disease. According to the proposed method, cases in Russia will start to decline continuously in the first week of June, and the cases will end completely in the last week of October. The cases reach a peak in Russia in May and June, according to our proposed method. The result of our proposed method shows that Russia will need some Hospital beds and ICU beds in May, June, and July when the number of cases is at a peak. The result shows the basic reproduction rate is 3.0 in starting, which starts decreasing visually after some time and goes up to one and even lower. A time-dependent SEAIHCRD model for Russia shows that the case fatality rate of COVID-19 in Russia is very low. There are many ups and downs in the daily case fatality rate with a maximum value of 1.6 in our model and then slows down, and total CFR is nearly 1.4. The proposed model for Russia showing that around 170^th^ day, the number of hospital beds and the number of ICU beds are required. That time the number of death cases is 300 per day, and daily-hospitalized cases are nearly 2000. The proposed method requires approximately 300 hospital beds and 60 ICU beds when the cases are at their peak in Russia. A time-dependent SEAIHCRD model results for Russia has shown in Fig. 20.

**Figure 20:**
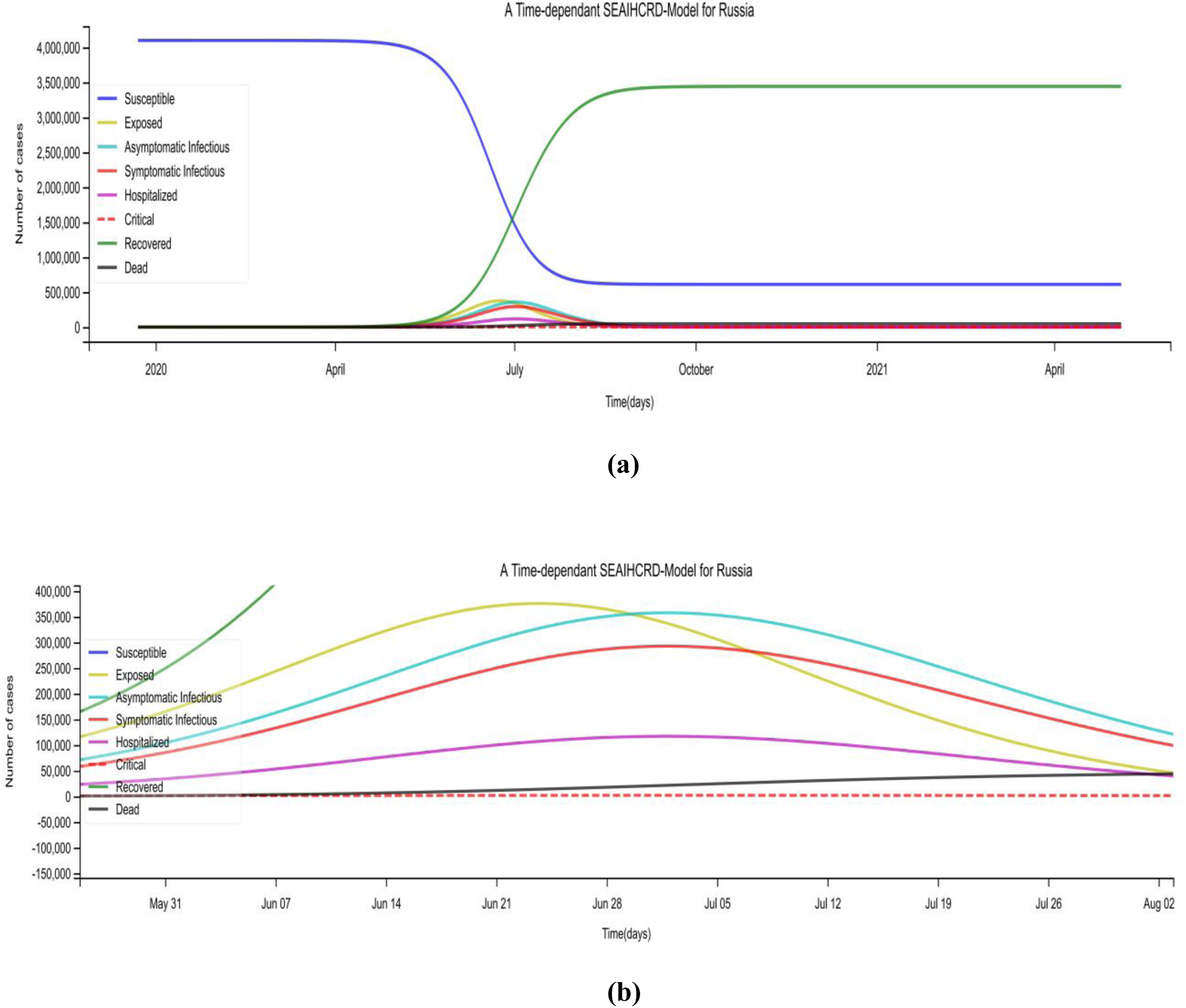
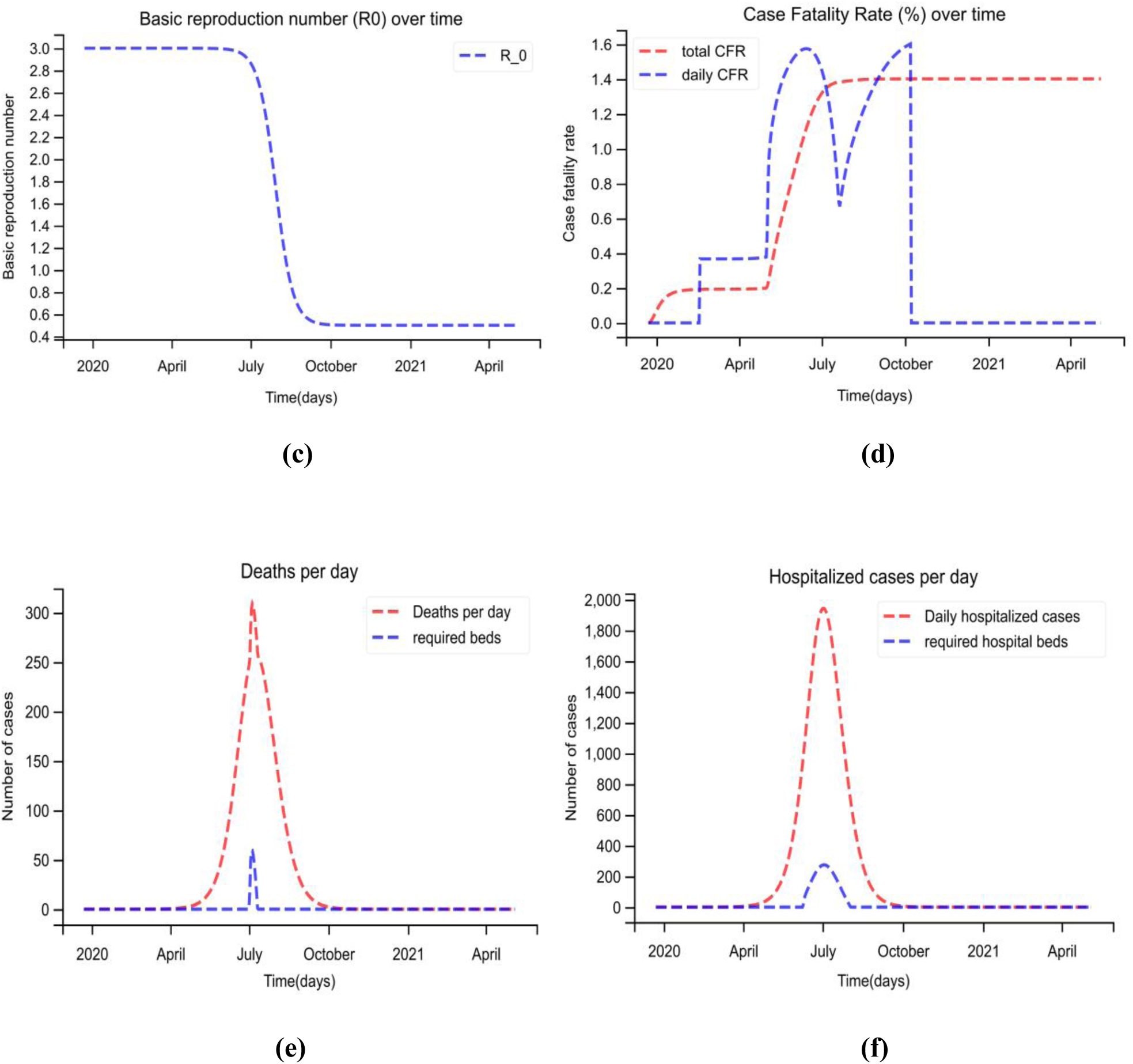
A time-dependent SEAIHCRD model for Russia. **(a)** The spread scenario of A time-dependent SEAIHCRD model for Russia. A 500-day analysis has been done by the proposed model, which starts from January 22. **(b)** In this, the peak point of the model has shown by zooming for cases. **(c)** The basic reproduction number of Russia over time **(d)** the case fatality rate of Russia over time. **(e)** Daily death cases in Russia, where the red line shows the number of deaths per day and the blue line shows how many ICU beds are required during peak days. **(f)** Russia’s Daily Hospital Cases, where the red line indicates the number of hospital cases per day, and the blue line shows how many hospital beds are required during peak days.

### 4.5 A time-dependent SEAIHCRD model for South Africa

South Africa is one of the most affected countries by COVID-19 disease. According to the proposed method, cases in South Africa will start to decline continuously in the third week of July, and the cases will end completely in the last week of October. The cases reach a peak in South Africa, according to our proposed method, in July and August. The result of our proposed method shows that South Africa will need some Hospital beds and ICU beds in July when the number of cases is at a peak. The result also indicates the basic reproduction rate is 4.5 in starting that decreases visually to values lower that one at a later time. A time-dependent SEAIHCRD model for South Africa shows that the case fatality rate of COVID-19 in South Africa is very low. The daily case fatality rate attains a maximum of 2.4 with many fluctuations in between and then slows down. The total CFR is coming out to be nearly 1.6. The proposed model for South Africa showing that around 175^th^ day, the number of hospital beds and the number of ICU beds are required. That time the number of death cases is 450 per day, and daily-hospitalized cases are nearly 1000. The proposed method requires approximately 150 hospital beds and 60 ICU beds when the cases are at their peak in South Africa. A time-dependent SEAIHCRD model results for South Africa has shown in Fig. 21.

**Figure 21:**
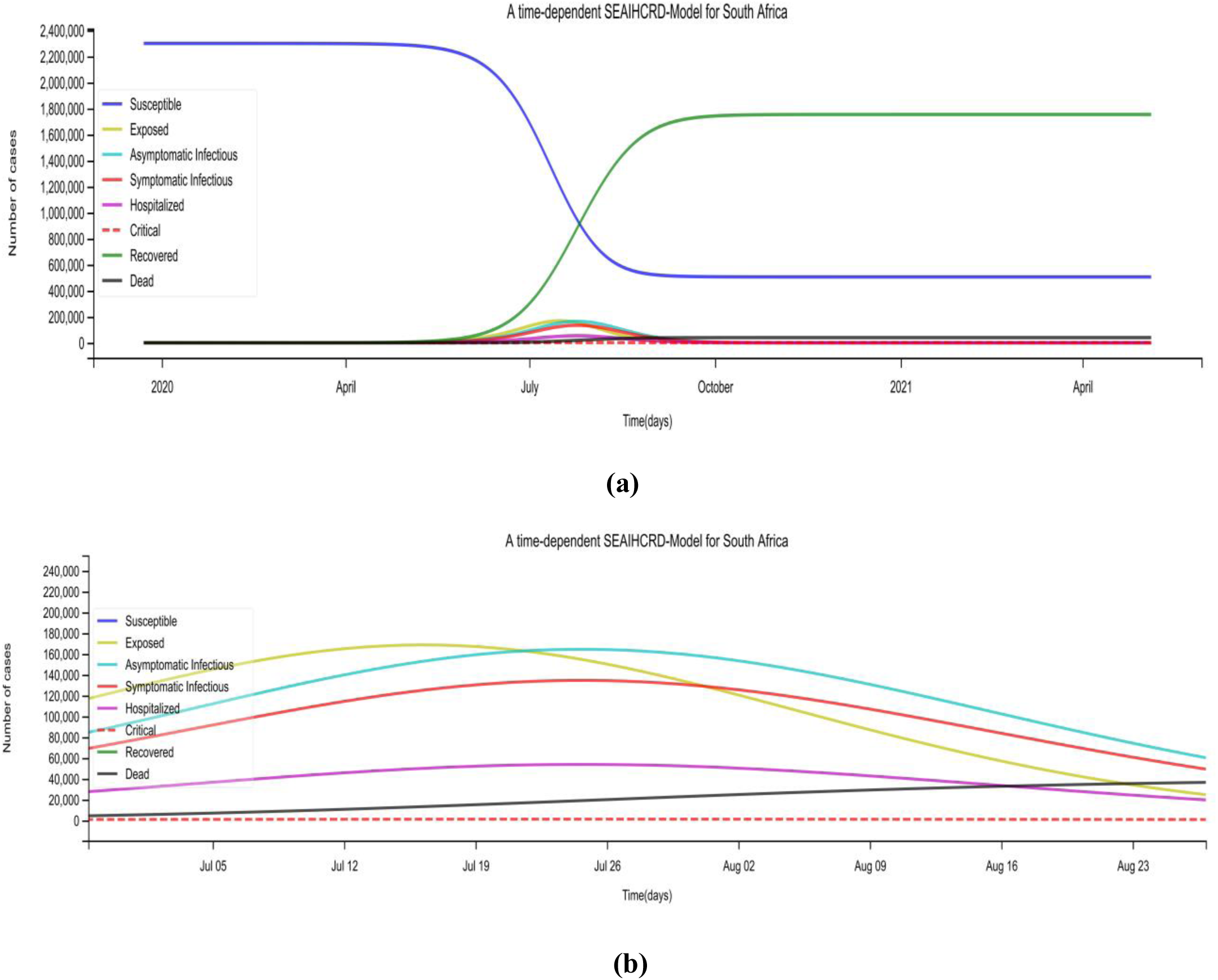
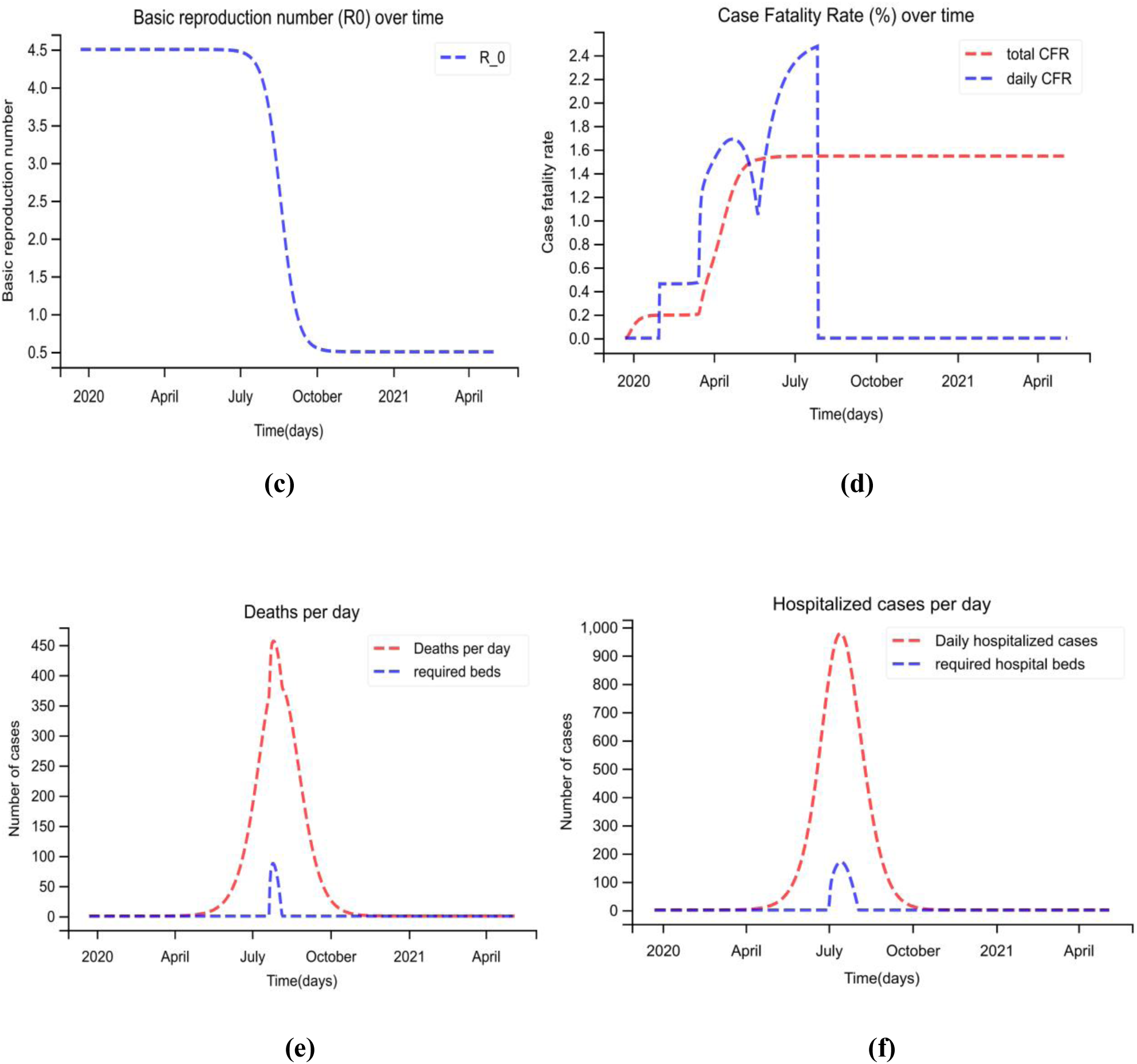
A time-dependent SEAIHCRD model for South Africa. **(a)** The spread scenario of A time-dependent SEAIHCRD model for South Africa. A 500-day analysis has been done by the proposed model, which starts from January 22. **(b)** In this, the peak point of the model has shown by zooming for cases. **(c)** The basic reproduction number of South Africa over time **(d)** the case fatality rate of South Africa over time. **(e)** Daily death cases in South Africa, where the red line shows the number of deaths per day and the blue line shows how many ICU beds are required during peak days. **(f)** South Africa’s Daily Hospital Cases, where the red line indicates the number of hospital cases per day, and the blue line shows how many hospital beds are required during peak days.

### 4.6 A time-dependent SEAIHCRD model for the United States

The United States is the most affected country by the COVID-19 disease. According to the proposed method, cases in the United States will start to decline continuously in the first week of July, and the cases will end completely in the last week of November. The cases reach on the peak in the United States, according to our proposed method, in July and August. The results of our proposed method show that the United States will need some Hospital beds and ICU beds in June, July, and August, when the number of cases is at a peak. The result shows the basic reproduction rate is 4.2 in starting, and after some time, it starts decreasing visually and goes up to one and even lower. A time-dependent SEAIHCRD model for the United States shows that the case fatality rate of COVID-19 in the United States is moderate. There are many ups and downs in the daily case fatality rate, but it goes higher and higher up to 5.5 in our model and then slows down, and total CFR is nearly 3.5. The proposed model for the United States is showing that around the 220^th^ day, the number of hospital beds and the number of ICU beds are required. That time the number of death cases is 3500 per day, and daily-hospitalized cases are nearly 14000. The proposed method requires approximately 1000 hospital beds and 250 ICU beds when the cases are at their peak in the United States. A time-dependent SEAIHCRD model results for the United States has shown in Fig. 22.

**Figure 22:**
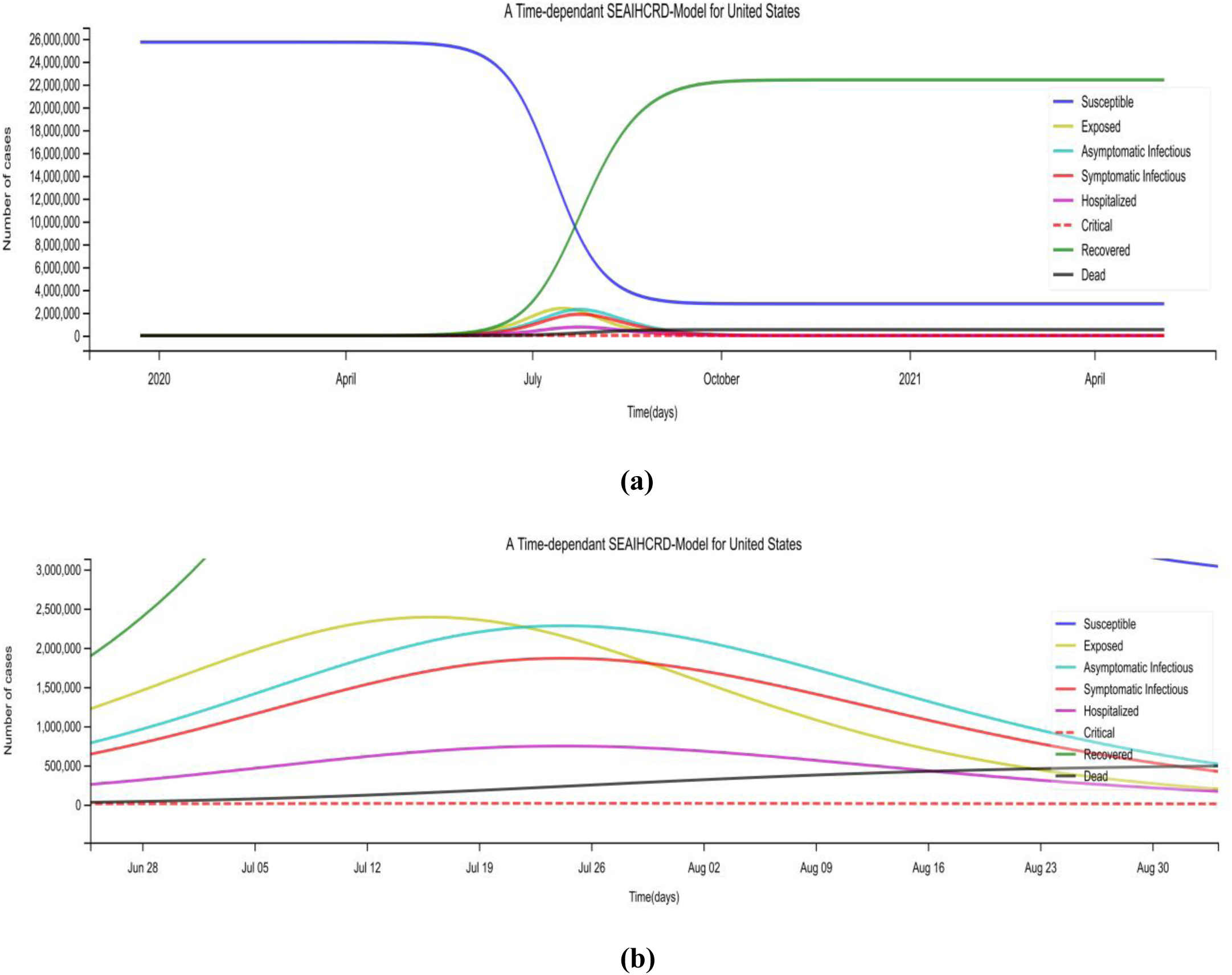
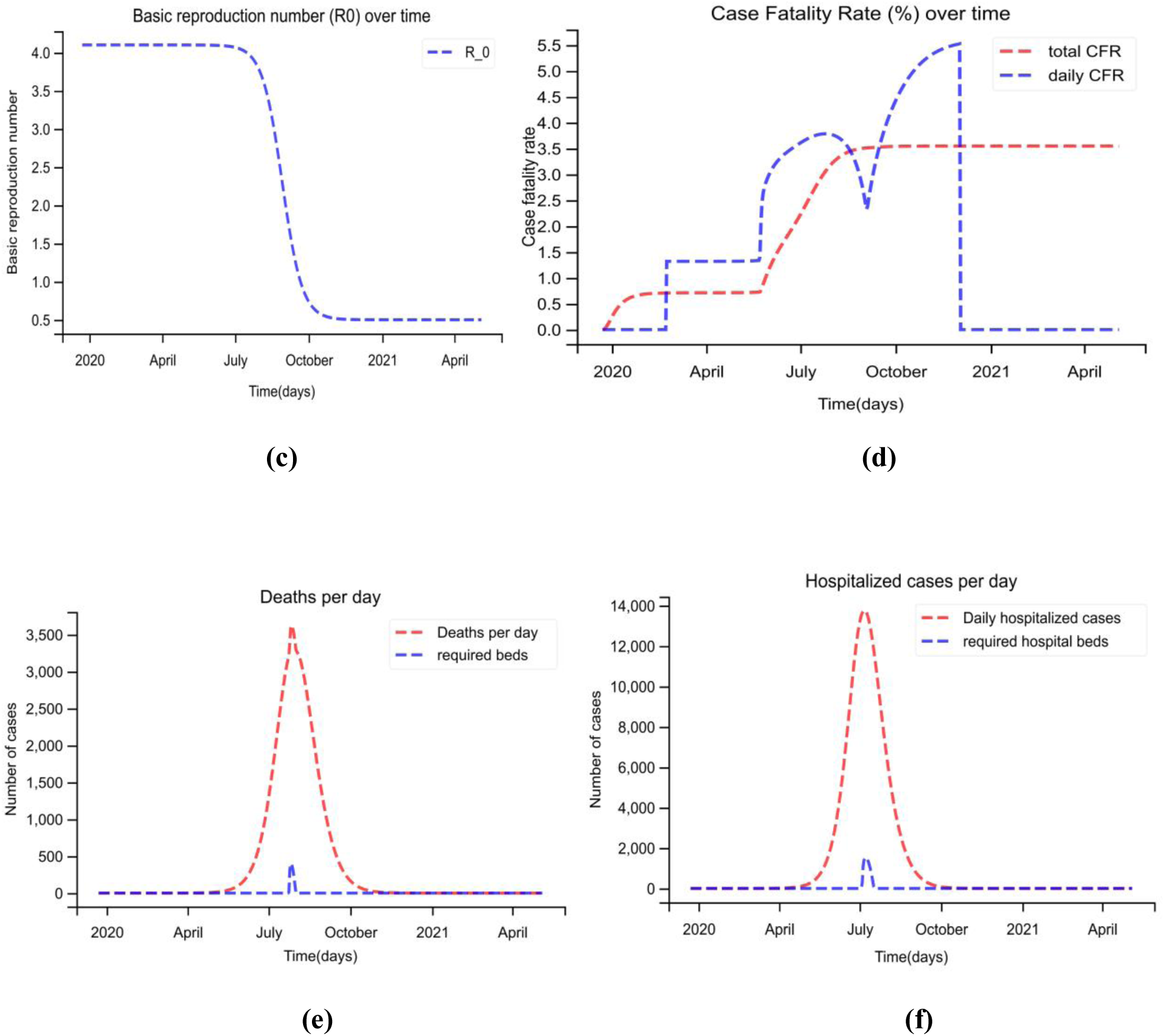
A time-dependent SEAIHCRD model for the United States. **(a)** The spread scenario of A time-dependent SEAIHCRD model for the United States. A 500-day analysis has been done by the proposed model, which starts from January 22. **(b)** In this, the peak point of the model has shown by zooming for cases. **(c)** The basic reproduction number of the United States over time **(d)** the case fatality rate of The United States over time. **(e)** Daily death cases in the United States, where the red line shows the number of deaths per day and the blue line shows how many ICU beds are required during peak days. **(f)** The United States Daily Hospital Cases, where the red line indicates the number of hospital cases per day, and the blue line shows how many hospital beds are required during peak days.

## Discussion

Results from the time-dependent SEAIHCRD model suggest that this model has excellent potential for predicting the incidence of COVID-19 diseases in time series. The central aspect of our proposed model is the addition of hospitalized, critical and asymptomatic infectious compartment, which improves the basic understanding of disease spread and results. Most of the cases are mild or asymptomatic, and asymptomatic cases do not show any symptoms. Still, after some time, some asymptomatic cases show disease symptoms; all of these scenarios are considered in the proposed model. We have the number of hospital beds and the number of ICU beds of the selected countries; based on that, we have calculated the number of beds required in the peak days of infection. Basic reproduction number and case fatality rate are the basic measures for any epidemic that we have also calculated. Sometimes conditions get worse because of a massive number of infected people at the same time, and the country does not have facilities to treat all critical cases when triage conditions occur. The time-dependent SEAIHCRD model can solve the problem of triage.

The model will propagate and forecast dynamic evolution after estimating the model parameters based on available clinical data. The proposed model is very sensitive to the initial conditions and values of the parameters. We are mainly focused on death cases because death cases hardly go undetected. The proposed model also estimates the approximate date when cases of death are unlikely to occur.

## Limitations

Modelling is one of the most powerful tools that give intuitive effects when multiple factors act together. No model is perfect. Like any other mathematical model, our proposed model also has some limitations as following

Our Ordinary Differential Equations system is very sensitive to initial parameters. Hence, while giving the initial parameters, one should be very careful. Small parameter changes can cause a massive difference in results. We put cases in the death compartment that are serious and not found treatment in ICU care. Our method is focused, in particular, on serious cases and deaths. The proposed model considers that the value of R_0_ cannot be increased; either it decreases or remains constant. We have assumed that the cases that have been recovered will be immunized, meaning that they will not be infected again.

## 5. Conclusion

A mathematical model is a potent tool for the quantitative behavior of any physical phenomenon, including any pandemic like COVID-19. Some mathematical models like the SIR model and SEIR model have been widely used for the prediction of disease outbreak and spread behavior. Our proposed time-dependent SEAIHCRD-Model is an extension of the SEIR-Model in which four-compartments are added that are Asymptomatic infectious, hospitalized, critical, and death compartment. Some people have not seen any symptoms of the disease. They are going to the asymptomatic infectious compartment. Some people have seen symptoms of the disease. They are going to the symptomatic infectious compartment. People with a serious infection who have severe pneumonia-like symptoms need hospitalization. These individuals may either recover or move to the critical compartment. The study indicates that most cases are mild or asymptomatic and are carefully monitored at home. Less percentage of people are critical, but if the patient is old or has some pre-existing disease, such as chronic respiratory disease, diabetes, cancer, hypertension, and cardiovascular disease, they are at high risk and may need treatment in an ICU. They either recover from the disease or die of it.

The proposed model estimates the date and magnitude of peaks of corresponding to exposed people, number of asymptomatic infectious people, symptomatic infectious people, the number of people hospitalized, number of people admitted in ICUs, and the number of death of COVID-19. This proposed model is time-dependent, so we calculate the number of cases asymptomatic infectious cases, symptomatic infectious cases, hospitalized cases, critical cases with time. We have COVID-19 data on hospital beds and ICU beds of most infected countries after we calculate the hospitalized cases and critical cases using the proposed model. Based on our calculation, we can tell how many beds are required. A time-dependent SEAIHCRD model also calculates the basic reproduction number over time, and its computational results are almost the same as the actual data, but sometimes vary. We also calculate the case fatality rate over time for the countries most affected by COVID-19. The proposed model calculates two types of Case fatality rate: one is CFR daily, and the other is total CFR.

According to the present time-dependent SEAIHCRD model, Mexico’s case fatality rate is very high, so the death cases are very high there, and with more critical cases, more ICU beds will be needed. The case fatality rate and basic reproduction number of India, South Africa, and Russia are quite low; hence, the number of deaths here is also low. The United States is most affected country by COVID-19 disease. The United States is taking more time to recover because of more number of cases there. The number of cases and death decreased in the country following the lock-down. Cases in India and Mexico will be at the top in August, and the number of hospital beds and ICU beds will be required during this peak period. India cases will almost stop coming in the first week of December. Brazil is one of the most affected countries by the COVID-19 disease just after the United States, and its cases will almost stop coming in the first week of November.

## Data Availability

We used public available data which are taken from trusted government agencies

https://www.who.int/

https://www.mohfw.gov.in/

## Acknowledgement

I would like to express my special thanks of gratitude to our collaborator Harel Dahari of The Program for Experimental and Theoretical Modeling, Division of Hepatology, Department of Medicine, Stritch School of Medicine, Loyola University Medical Center, Maywood IL, United States and as well as Jonathan Ozik from Consortium for Advanced Science and Engineering, University of Chicago, Chicago, IL, USA, who gave me the golden opportunity to do work with them on SPARC project that helped us in doing a lot of good Research and we came to know about so many new things and I am really thankful to them.

The work has been supported by a grant received from the Ministry of Education, Government of India under the Scheme for the Promotion of Academic and Research Collaboration (SPARC) (ID: SPARC/2019/1396).

